# Risk of SARS-CoV-2 reinfection during multiple Omicron variant waves in the UK general population

**DOI:** 10.1101/2023.06.29.23292043

**Authors:** Jia Wei, Nicole Stoesser, Philippa C. Matthews, Tarnjit Khera, Owen Gethings, Ian Diamond, Ruth Studley, Nick Taylor, Tim E. A. Peto, A. Sarah Walker, Koen B. Pouwels, David W. Eyre, COVID-19 Infection Survey team

## Abstract

SARS-CoV-2 reinfections increased substantially after Omicron variants emerged. Large-scale community-based comparisons across multiple Omicron waves of reinfection characteristics, risk factors, and protection afforded by previous infection and vaccination, are limited, especially after widespread national testing stopped. We studied 245,895 adults ≥18y in the UK’s national COVID-19 Infection Survey with at least one infection (identified from positive swab tests done within the study, linked from national testing programmes, or self-reported by participants, up to their last study assessment). We quantified the risk of reinfection in multiple infection waves, including those driven by BA.1, BA.2, BA.4/5, and most recently BQ.1/CH.1.1/XBB.1.5 variants, in which most reinfections occurred. Reinfections had higher cycle threshold (Ct) values (lower viral load) and lower percentages self-reporting symptoms compared with first infections. Across multiple Omicron waves, protection against reinfection was significantly higher in those previously infected with more recent than earlier variants, even at the same time from previous infection. Protection against Omicron reinfections decreased over time from the most recent infection if this was the previous or penultimate variant (generally within the preceding year), but did not change or even slightly increased over time if this was with an even earlier variant (generally >1 year previously). Those 14-180 days after receiving their most recent vaccination had a lower risk of reinfection with all Omicron variants except BA.2 than those >180 days from their most recent vaccination. Reinfection risk was independently higher in those aged 30-45 years, and with either low or high Ct values in their most recent previous infection. Overall, the risk of Omicron reinfection is high, but with lower severity than first infections; reinfection risk is likely driven as much by viral evolution as waning immunity.

## Introduction

Omicron (B.1.1.529) became the dominant severe acute respiratory syndrome coronavirus 2 (SARS-CoV-2) variant globally in December 2021. Due to a high number of mutations in the viral spike protein, it has enhanced transmissibility and infectivity compared with previous variants and can more easily escape immunity acquired from both previous infection and vaccination^1–7^, causing widespread infection worldwide. The Omicron variant has further mutated and recombined into different subvariants, which have caused multiple infection waves. In the UK, there have been five Omicron infection waves from December 2021 to January 2023, caused by the dominant subvariants BA.1, BA.2, BA.4/5, BQ.1 (a sub-lineage of BA.4/5) and most recently a mixture of BQ.1 and BA.2.75 and its sub-lineages, particularly CH.1 and XBB.1.5^8^.

SARS-CoV-2 reinfections have been reported since mid 2020^9–11^. Before the emergence of the Omicron variant, SARS-CoV-2 reinfections were relatively uncommon^12,13^, and previous infections provided 80-90% protection against a pre-Omicron reinfection^14,15^. However, reinfection risk increased substantially with Omicron^15–17^, with the protection against an Omicron BA.1 and BA.2 reinfection from a previous non-Omicron infection estimated to be only 20-45%^18–20^. Reinfections with different Omicron subvariants also occur, although protection from a previous Omicron infection against a new Omicron reinfection has been found to be greater at 50-90%^18,21^. However, rapid waning of protection against a new BA.5 infection following BA.1/BA.2 infection has also been reported^22^.

Understanding the extent and duration of protection against reinfection associated with vaccination and prior infection is important for planning vaccination and other control measures. However, a major challenge is that much information about infection comes from routine testing programmes, which may have numerous biases in terms of who decides to test, access to testing (particularly since tests are no longer free for most of the population in many countries, such as the UK), and over-representation of symptomatic infections, leading to incomplete ascertainment of infection history and potential bias in estimating reinfection severity, risk factors, protection from previous infection and vaccination, especially for the most recent Omicron infection waves after testing programmes were discontinued, as in the UK from 1 April 2022.

We therefore used data from the UK’s Office for National Statistics (ONS) COVID-19 Infection Survey (CIS), which undertakes regular longitudinal testing of participants, linked to data from the English and Welsh national testing programmes and supplemented by self-reported positive swab tests, to examine the characteristics and severity of SARS-CoV-2 reinfections compared with first infections. We used flexible parametric survival models to examine the risk factors for reinfections in multiple Omicron waves of the pandemic including those driven by BA.1, BA.2, and BA.4/5 variants, and the most recent wave with a mixture of BQ.1, CH.1, and XBB1.5 subvariants, and specifically investigated the protective effects from previous vaccination and infection.

## Results

From 28 February 2020 to 13 March 2023, 245,895 participants ≥18y were infected with SARS-CoV-2 based on a positive swab test in the study, national testing programmes or self-reported at any time up to their final study assessment (see **Methods**; self-report only in 43,246 (17.6%)). 45,137 (18%) were reinfected; 3,513 (1.4%) participants had three infections, 162 (0.07%) four, and 7 five infections (**Table S1, Figure S1**). All of the seven participants identified as having been infected 5 times were white females, aged 22-49 years. Three reported having a long-term health condition, and two were healthcare workers. The variants, symptoms, and Ct values of these infections were shown in **Table S1**.

The median age at first infection was 53 years, and 48 and 45 years at the first and second reinfection (i.e. second and third infection), respectively (**Table 1**). 54.6% of those ever infected were female, versus 58.0% and 64.8% among those reinfected once or twice. 93.4% of those ever infected reported white ethnicity, more than those reinfected once (92.5%) or twice (90.8%). Those reporting working in healthcare were more likely to have reinfections, accounting for 5.3%, 6.9%, 9.0%, and 9.3% of those who had one, two, three, and four infections, versus 4.4% of the cohort as a whole.

**Table 1.**
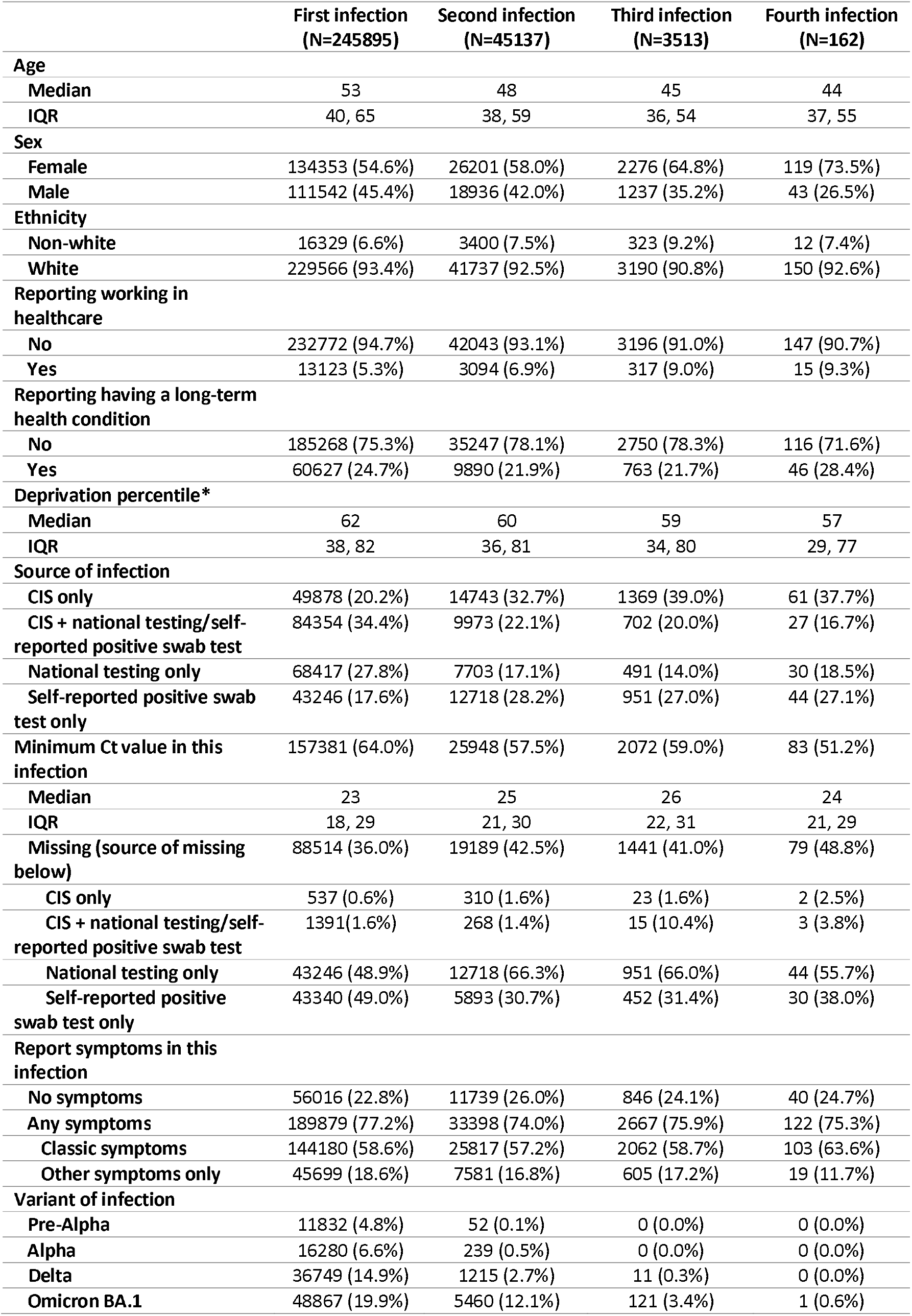
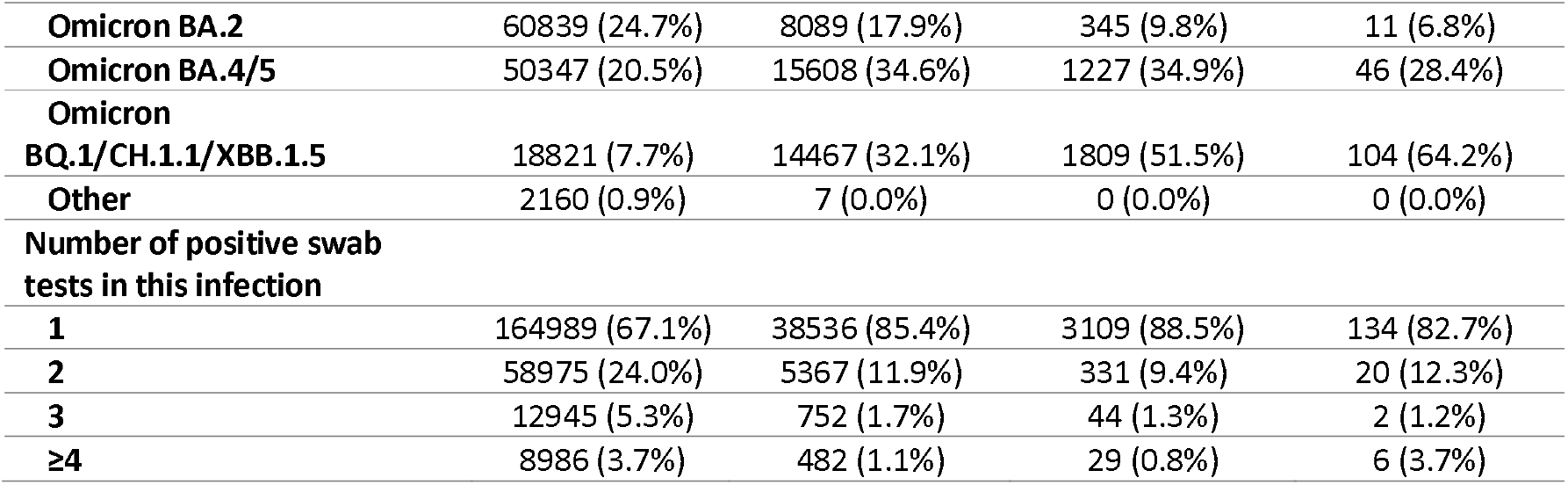
Characteristics of participants who had a first, second, third, and fourth infection. *A higher deprivation percentile represents less deprived. CIS=COVID-19 Infection Survey. Infections were identified from positive swab tests done within the study, linked from national testing programmes, or self-reported by participants (participants were asked not to self-report study tests). Symptoms were determined by all symptoms from the list below reported by participants within [0,35] days of the first positive test in each infection, including symptoms reported in the last 7 days of study assessments and symptoms reported when participants thought they had COVID. Symptoms were classified as ‘classic’ (any of cough, fever, loss of taste/smell) or ‘other’ (myalgia, fatigue/weakness, sore throat, shortness of breath, headache, diarrhoea, nausea, abdominal pain). If no symptoms were reported within CIS, we included any self-reported symptoms available from national testing programmes as “other” as specific symptoms were not available from this data source. Variant was defined by sequencing data where available, otherwise by S-gene presence/absence and calendar time reflecting periods when specific variants dominated in the UK (see Methods). In all, 460,026 participants ≥18y were ever swabbed in the survey, median (IQR) age at first swab was 54 (39-67) years, 53.6% were female, 7.0% reported non-white ethnicity, 4.4% reported working in healthcare and 26.8% a long-term health condition.

Main results
|  | First infection<br>(N=245895) | Second infection<br>(N=45137) | Third infection<br>(N=3513) | Fourth infection<br>(N=162) |
| --- | --- | --- | --- | --- |
| **Age** |  |  |  |  |
| Median | 53 | 48 | 45 | 44 |
| IQR | 40, 65 | 38, 59 | 36, 54 | 37, 55 |
| **Sex** |  |  |  |  |
| Female | 134353 (54.6%) | 26201 (58.0%) | 2276 (64.8%) | 119 (73.5%) |
| Male | 111542 (45.4%) | 18936 (42.0%) | 1237 (35.2%) | 43 (26.5%) |
| **Ethnicity** |  |  |  |  |
| Non-white | 16329 (6.6%) | 3400 (7.5%) | 323 (9.2%) | 12 (7.4%) |
| White | 229566 (93.4%) | 41737 (92.5%) | 3190 (90.8%) | 150 (92.6%) |
| **Reporting working in healthcare** |  |  |  |  |
| No | 232772 (94.7%) | 42043 (93.1%) | 3196 (91.0%) | 147 (90.7%) |
| Yes | 13123 (5.3%) | 3094 (6.9%) | 317 (9.0%) | 15 (9.3%) |
| **Reporting having a long-term health condition** |  |  |  |  |
| No | 185268 (75.3%) | 35247 (78.1%) | 2750 (78.3%) | 116 (71.6%) |
| Yes | 60627 (24.7%) | 9890 (21.9%) | 763 (21.7%) | 46 (28.4%) |
| **Deprivation percentile\*** |  |  |  |  |
| Median | 62 | 60 | 59 | 57 |
| IQR | 38, 82 | 36, 81 | 34, 80 | 29, 77 |
| **Source of infection** |  |  |  |  |
| CIS only | 49878 (20.2%) | 14743 (32.7%) | 1369 (39.0%) | 61 (37.7%) |
| CIS + national testing/self-reported positive swab test | 84354 (34.4%) | 9973 (22.1%) | 702 (20.0%) | 27 (16.7%) |
| National testing only | 68417 (27.8%) | 7703 (17.1%) | 491 (14.0%) | 30 (18.5%) |
| Self-reported positive swab test only | 43246 (17.6%) | 12718 (28.2%) | 951 (27.0%) | 44 (27.1%) |
| Minimum Ct value in this infection | 157381 (64.0%) | 25948 (57.5%) | 2072 (59.0%) | 83 (51.2%) |
| Median | 23 | 25 | 26 | 24 |
| IQR | 18, 29 | 21, 30 | 22, 31 | 21, 29 |
| Missing (source of missing below) | 88514 (36.0%) | 19189 (42.5%) | 1441 (41.0%) | 79 (48.8%) |
| CIS only | 537 (0.6%) | 310 (1.6%) | 23 (1.6%) | 2 (2.5%) |
| CIS + national testing/self-reported positive swab test | 1391 (1.6%) | 268 (1.4%) | 15 (10.4%) | 3 (3.8%) |
| National testing only | 43246 (48.9%) | 12718 (66.3%) | 951 (66.0%) | 44 (55.7%) |
| Self-reported positive swab test only | 43340 (49.0%) | 5893 (30.7%) | 452 (31.4%) | 30 (38.0%) |
| **Report symptoms in this infection** |  |  |  |  |
| No symptoms | 56016 (22.8%) | 11739 (26.0%) | 846 (24.1%) | 40 (24.7%) |
| Any symptoms | 189879 (77.2%) | 33398 (74.0%) | 2667 (75.9%) | 122 (75.3%) |
| Classic symptoms | 144180 (58.6%) | 25817 (57.2%) | 2062 (58.7%) | 103 (63.6%) |
| Other symptoms only | 45699 (18.6%) | 7581 (16.8%) | 605 (17.2%) | 19 (11.7%) |
| **Variant of infection** |  |  |  |  |
| Pre-Alpha | 11832 (4.8%) | 52 (0.1%) | 0 (0.0%) | 0 (0.0%) |
| Alpha | 16280 (6.6%) | 239 (0.5%) | 0 (0.0%) | 0 (0.0%) |
| Delta | 36749 (14.9%) | 1215 (2.7%) | 11 (0.3%) | 0 (0.0%) |
| Omicron BA.1 | 48867 (19.9%) | 5460 (12.1%) | 121 (3.4%) | 1 (0.6%) |
| **Omicron BA.2** | 60839 (24.7%) | 8089 (17.9%) | 345 (9.8%) | 11 (6.8%) |
| **Omicron BA.4/5** | 50347 (20.5%) | 15608 (34.6%) | 1227 (34.9%) | 46 (28.4%) |
| **Omicron BQ.1/CH.1.1/XBB.1.5** | 18821 (7.7%) | 14467 (32.1%) | 1809 (51.5%) | 104 (64.2%) |
| **Other** | 2160 (0.9%) | 7 (0.0%) | 0 (0.0%) | 0 (0.0%) |
| **Number of positive swab tests in this infection** |  |  |  |  |
| **1** | 164989 (67.1%) | 38536 (85.4%) | 3109 (88.5%) | 134 (82.7%) |
| **2** | 58975 (24.0%) | 5367 (11.9%) | 331 (9.4%) | 20 (12.3%) |
| **3** | 12945 (5.3%) | 752 (1.7%) | 44 (1.3%) | 2 (1.2%) |
| **≥4** | 8986 (3.7%) | 482 (1.1%) | 29 (0.8%) | 6 (3.7%) |

38.9% and 39.1% of participants whose first infection was with Pre-Alpha or Alpha variants had a second infection identified, with BA.1 and BA.2 being the most common reinfection variant followed by BA.4/5. 35.6% of participants whose first infection was with Delta variants had a second infection, among which 35.1% were BA.4/5 (**Figure S1**).

Using Kaplan-Meier estimation from the start of the earlier infection, 50% of participants were reinfected by 799 days (95% Confidence Interval [CI] 786-820) if the earlier infection was with Pre-Alpha variants, versus 733 days (95% CI 719-755) with Alpha variants, and 601 days (591-610) with Delta variants. 25% of participants were reinfected by 521 (516-528), 452 (448-457), 326 (322-331), 368 (364-371), and 390 (382-394) days if the earlier infection was with Pre-Alpha, Alpha, Delta, BA.1, BA.2 variants, respectively (**Figure S2**).

### Cycle threshold (Ct) values and reinfections

185,484 (62.9%) infections had a Ct value recorded from the same TaqPath assay (see Methods). Missing Ct values were largely from infections identified from national testing only and not using this test (45.5% of the missing Ct) and self-report only (52.1% of the missing Ct). The median observed Ct was 23 in first, 25 in second, and 26 in third infections (p<0.001) (**Table 1**).

Using robust linear regression, independently, and allowing for interactions between infection number, variant, and time from vaccination, Ct values were higher in reinfections than first infections across different variants, excepting only reinfections with Omicron subvariants in those not vaccinated (**Figure 1**). Ct values were lower in males (p<0.001), healthcare workers (p=0.001), those who had a higher deprivation score (p<0.001), those reporting symptoms (p<0.001), and in participants with a higher number of positive tests within infection episodes (p<0.001) (**Table S2**). Compared with BA.1 variant infections, Pre-Alpha and Alpha variant infections had higher mean Ct values, while Delta had lower mean Ct values (p<0.001). Among different Omicron subvariants, BA.2, BA.4/5, and BQ.1/CH.1.1/XBB.1.5 infections all had progressively higher mean Ct value than BA.1 infections (p<0.001). Being 14-180 days from the most recent vaccination was independently associated with higher Ct values, and there were no substantial differences in this effect across variants and infections, considering interactions.

**Figure 1.**
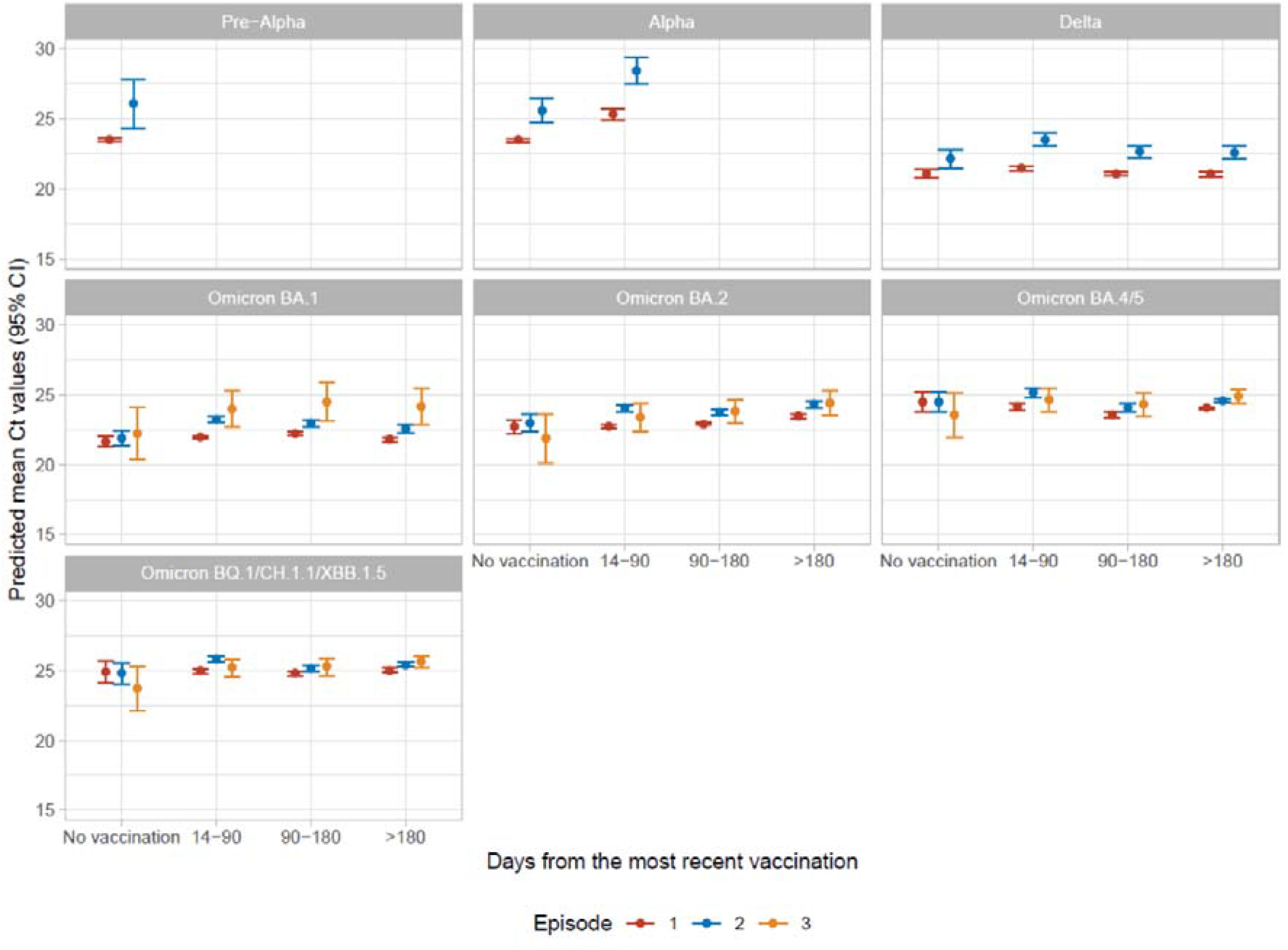
Predicted mean Ct values (95% CIs) by infection variant, days from the most recent vaccination, and infection (first, second, or third). The 95% CIs are calculated as predictions ± 1.96 × standard error of the predictions. Predictions are plotted at the reference value of other variables (age=40 years, female, white ethnicity, not reporting working in healthcare, not reporting having a long-term health condition, deprivation percentile=60, reporting classic symptoms, having one positive test in the infection (estimates shown in **Table S2**).

In a separate model only including reinfections, either lower or higher Ct values in the most recent previous infection were associated with lower Ct values in the current reinfection (p<0.001) (**Figure S3**), while reporting classic symptoms (any of fever, cough, loss of smell, loss of taste) in the most recent previous infection was associated with higher Ct values in current reinfection (p=0.04) (**Table S2**).

### Symptoms and reinfections

The unadjusted percentage reporting any symptoms was slightly lower in the second infection (74.0%) and the third infection (75.9%) compared with the first infection (77.2%) (p<0.001). The second infection had a slightly lower percentage reporting classic symptoms than the first infection (57.2% vs 58.6%, p<0.001), but there was no evidence of differences with the third and fourth infections. However, the unadjusted percentages reporting only ‘other’ symptoms (myalgia, fatigue/weakness, sore throat, shortness of breath, headache, diarrhoea, nausea, abdominal pain) were higher in the first infection than the first, second and third reinfection (18.6% vs 16.8%, 17.2%, 11.7% respectively, p<0.001) (**Table 1**).

Using a multinomial logistic regression model, adjusting for multiple other factors including age, sex, ethnicity, reporting healthcare work, reporting long-term health conditions, social deprivation, time from most recent vaccination, and infection variant, classic symptoms were less commonly reported in a first reinfection than a first infection (relative risks RR=0.59 [95%CI 0.57-0.60]), as were other symptoms (RR=0.88 [0.85-0.91]). Classic symptoms were even less commonly reported in a second reinfection than a first infection (RR=0.52 [0.48-0.57]), but other symptoms were not (RR=1.02 [0.92-1.14]). Results remained similar after additionally adjusting for Ct values of the current infection, with slightly lower RRs of reporting classic symptoms in reinfections than a first infection (RR=0.52 [0.50-0.54], 0.46 [0.41-0.51] for first and second reinfection vs first infection) (**Table 2**).

**Table 2.** The association (relative risks [RR], 95% confidence intervals (95%CIs)) between reported classic or other symptoms and reinfections. . Multivariable multinomial regression model was adjusted for age, sex, ethnicity, reporting working in healthcare, reporting having a long-term health condition, deprivation percentile, infection variant, time from most recent vaccination. A separate model was built further adjusted for Ct values in the current infection. The 95% CIs are calculated as estimates ± 1.96 × standard error of the estimates.

|  | No symptom | Classic symptoms |  |  | Other symptoms |  |  |
| --- | --- | --- | --- | --- | --- | --- | --- |
| Model without Ct |  |  |  |  |  |  |  |
|  | reference | RR | 95%CI | p-value | RR | 95%CI | p-value |
| Infection (2 vs 1) |  | 0.52 | 0.50-0.54 | <0.001 | 0.84 | 0.80-0.88 | <0.001 |
| Infection (3 vs 1) |  | 0.46 | 0.41-0.51 | <0.001 | 1.03 | 0.83-1.10 | 0.5 |
| Model adjusted for Ct |  |  |  |  |  |  |  |
|  | reference | RR | 95%CI | p-value | RR | 95%CI | p-value |
| Infection (2 vs 1) |  | 0.59 | 0.57-0.60 | <0.001 | 0.88 | 0.85-0.91 | <0.001 |
| Infection (3 vs 1) |  | 0.52 | 0.48-0.57 | <0.001 | 1.02 | 0.92-1.14 | 0.7 |

### The risk of reinfection in multiple Omicron waves

42,582, 83,382, 164,263, and 184,566 adults ≥18y had had a previous infection and were at risk of reinfection for all or part of time periods where a single variant dominated in the UK, specifically the Omicron BA.1 (27 December 2021 to 6 February 2022), BA.2 (14 March 2022 to 22 May 2022), BA.4/5 (27 June 2022 to 6 November 2022), and BQ.1/CH.1.1/XBB.1.5 (6 November 2022 to 13 March 2023), respectively (denoted “waves”). During these periods, new infections could be most confidently assigned to specific variants, even in the absence of sequencing or S-gene presence/absence (see Methods). 3,504, 5,644, 15,079, and 16,076 participants had an Omicron BA.1, BA.2, BA.4/5, or BQ.1/CH.1.1/XBB.1 reinfection, and the Kaplan-Meier reinfection percentages were 10%, 11%, 14%, and 16% at the end of each wave, respectively, with corresponding reinfection incidences 24, 14, 10, 10 per 10,000 participant days, respectively. The unadjusted reinfection rate started at very high levels then decreased over time in BA.1 and BA.2 waves, remained lower throughout the BA.4/5 wave, and increased again in BQ.1/CH.1.1/XBB.1 wave, suggesting that the risk was shifting antigenically (**Figure S4**).

Across waves, adjusting for multiple confounders, protection against reinfection was higher following previous infection with more recent variants than earlier variants, including when the time from previous infection overlapped between variants (**Figure 2)**. There were variable effects of time since previous infection on risk of reinfection, suggesting that, rather than solely being driven by waning over time, changes in variant were a major driver of reinfection risk in most waves. In particular, consistently across all Omicron waves, protection against reinfections decreased over time from the most recent infection if this was the previous (thick solid lines) or penultimate (thin solid lines) variant (generally reflecting the most recent prior infection being within the preceding year), but did not change or even slightly increased over time if this most recent prior infection was with an even earlier variant (dashed lines) (generally more than a year previously). For example, there was no clear evidence that the protection arising from previous Pre-Alpha and Alpha infections changed over time from infection in the BA.1, BA.2, and BA.4/5 waves (p=0.9, 0.4 in BA.1, 0.8, 0.9 in BA.2, 0.4, 0.1 in BA.4/5), and somewhat counter-intuitively protection arising from previous Alpha infections increased over time in the BQ.1/CH.1.1/XBB.1.5 wave (Alpha: HR=0.86 per 60 days, p=0.01, noting that these participants had already avoided reinfection with all variants from Delta onwards). Similarly, protection from a previous Delta infection against a reinfection in the BA.2 wave gradually declined over time (reinfection HR=1.14 per 60 days, p<0.001), but did not change over time in the BA.4/5 wave (p=0.9), and increased over time in the BQ.1/CH.1.1/XBB.1.5 wave (HR=0.89 per 60 days, p<0.001; again these participants had avoided reinfection with all previous Omicron variants). Clearer waning of protection over time was seen after Omicron infections, for example, in the BA.4/5 wave, protection decreased over time from previous BA.1 (HR=1.17 per 60 days, p<0.001) and BA.2 infections (HR=1.31 per 60 days, p<0.001). In BQ.1/CH.1.1/XBB.1.5 wave, the protection was highest after a previous BA.4/5 infection, followed by a previous BA.2 infection, but protection also waned over time from infection (BA.2: HR=1.18 per 60 days, p<0.001, BA.4/5: HR=1.44 per 60 days, p<0.001). Results remained similar when modelling time from previous infection categorically (**Figure S7**).

**Figure 2.**
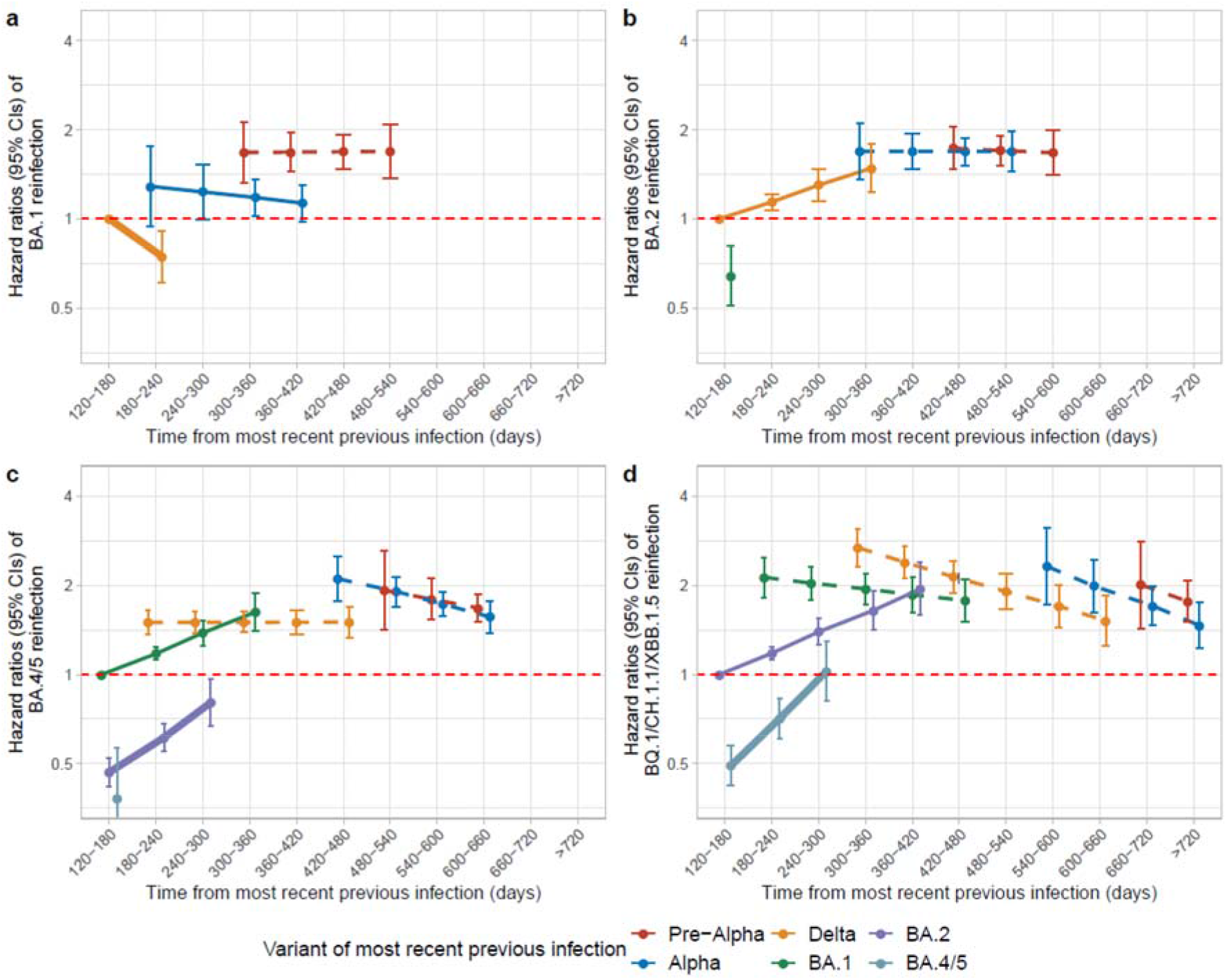
Risk (Hazard Ratios with 95% CIs) of reinfections during the Omicron BA.1 (a), Omicron BA.2 (b), Omicron BA.4/5 (c) and Omicron BQ.1/CH.1.1/XBB.1.5 (d) waves by time from most recent previous infection and variant of most recent previous infection. Time from previous infection was categorised as 120-180, 180-240, 240-300, 300-360, 360-420, 420-480, 480-540, 540-600, 600-660, 660-720, >720 days and its effect modelled as a trend over these categories (see **Figure S7** for categorical effects). Risk is presented versus a reference category of 120-180 days from an infection in the wave starting ∼6 months before the current wave (Delta for BA.1 and BA.2 waves, BA.1 for BA.4/5 waves and BA.2 for BQ.1/CH.1.1/XBB.1.5 waves). Line type and width represent the sequence of variants for better comparisons across waves (thick solid line represents the previous variant, thin solid line represents the penultimate variant, and dashed lines represent earlier variants). The 95% CIs are calculated as exponent of estimates ± 1.96 × standard error of the estimates. Adjusted (**Table S3**) for age, sex, ethnicity, reporting working in healthcare, reporting having a long-term health condition, deprivation percentile, infection variant, time from most recent vaccination (Figure 4), region, number of previous infections, symptoms in most recent infection and whether any previous infection had Ct<30 or was LFD positive. Results remain similar in sensitivity analyses adjusted for background infection prevalence (**Table S4**).

Time from the most recent vaccination was independently associated with the risk of reinfection. Compared with being >180 days from the most recent vaccination, the risk of reinfection was significantly lower 14-90 days and 90-180 days from the most recent vaccination in BA.1, BA.4/5, and BQ.1/CH.1.1/XBB.1.5 waves, but there was no evidence of difference in the BA.2 wave, although estimates were numerically lower (**Figure 3**).

**Figure 3.**
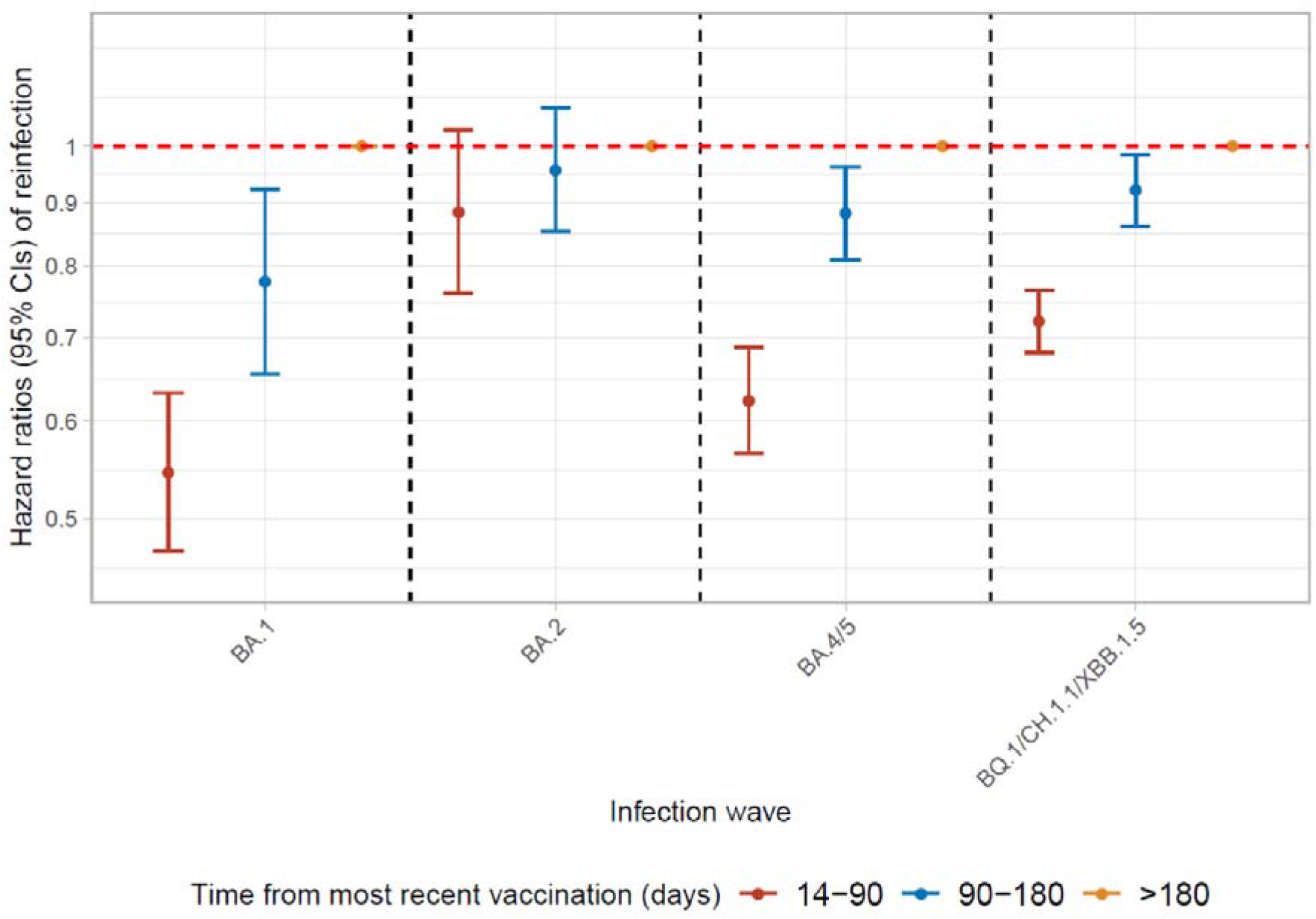
Risk (Hazard Ratios with 95% CIs) of Omicron reinfections in different waves by time from most recent vaccination. Time from most recent vaccination was split into 14-90 days, 90-180 days, >180 days. >180 days was used as reference category. Bivalent vaccines were only used from September 2022 onwards, and therefore would only have been relevant to infections in the final wave. The 95% CIs are calculated by estimates ± 1.96 × standard error of the estimates. Adjusted (**Table S3**) for age, sex, ethnicity, reporting working in healthcare, reporting having a long-term health condition, deprivation percentile, infection variant, time from most recent infection (Figure 3), region, number of previous infections, symptoms in most recent infection and whether any previous infection had Ct<30 or was LFD positive. Results remain similar in sensitivity analyses adjusted for background infection prevalence (**Table S4**).

Across waves, reinfection risk was consistently higher in young to middle-aged participants (30-45y) (**Figure S5**), females, and those reporting white ethnicity, working in healthcare, and long-term health conditions (**Table S3**). Reporting classic symptoms in the most recent previous infection was associated with a lower risk of reinfection in BA.1, BA.2 waves, but a higher risk in the BQ.1/CH.1.1/XBB.1.5 wave. Participants living in Northern Ireland, Scotland, and Wales had a lower risk of BA.1 and BA.2 reinfection, but those living in Northern Ireland and Wales had a higher risk of BQ.1/CH.1.1/XBB.1.5 reinfection (**Figure 4)**. Reinfection risk was consistently higher in those with an intermediate Ct value (20-30) in previous infection across all waves (**Figure S6**).

**Figure 4.**
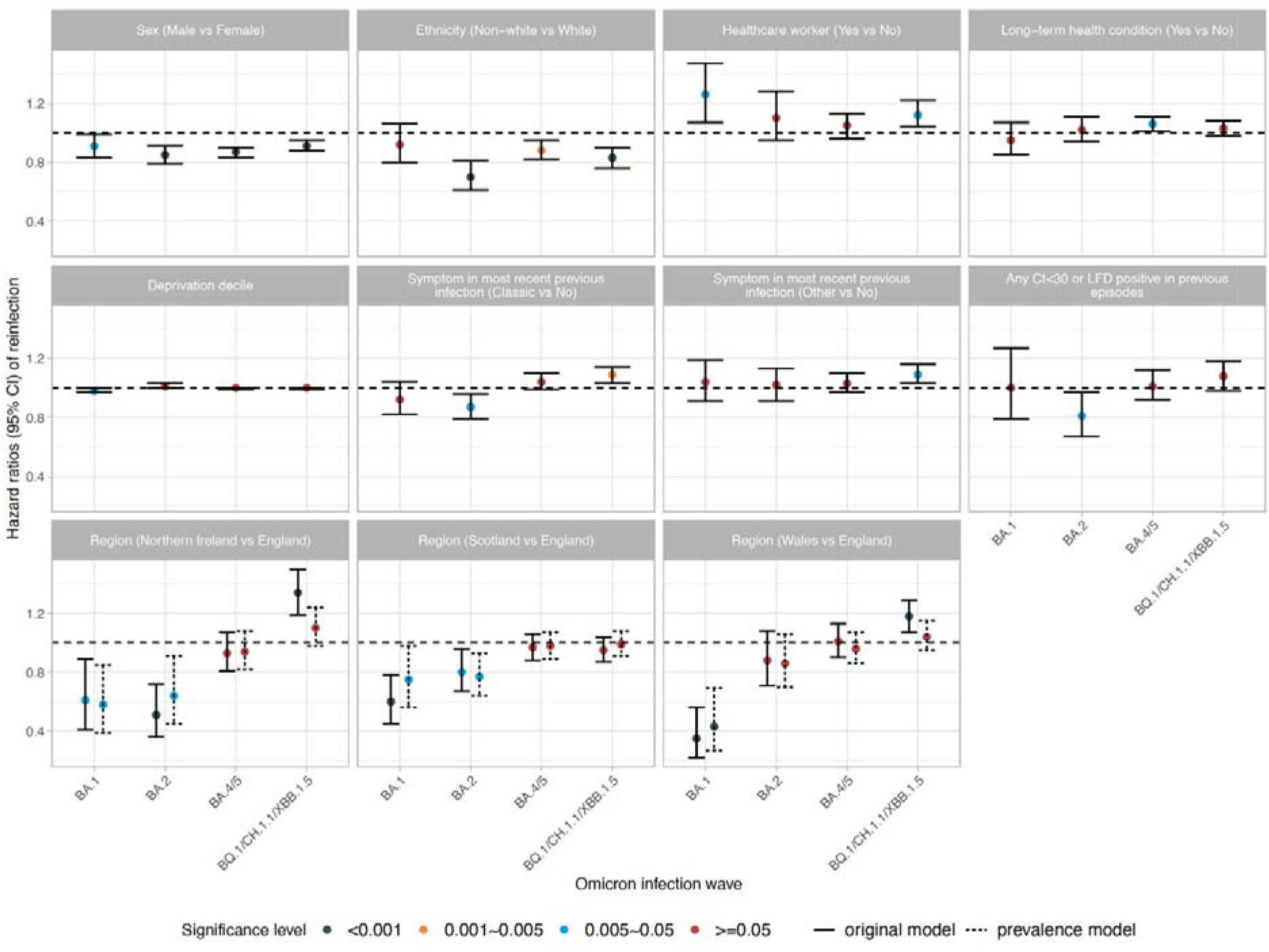
The association (hazard ratios, 95%CI) between different covariates and the risk of reinfection across multiple Omicron infection waves (BA.1, BA.2, BA.4/5, BQ.1/CH.1.1/XBB.1.5). Estimates shown in **Table S3**. Results remain similar in sensitivity analyses adjusted for background infection prevalence (**Table S4**). Small differences in associations with region between two models are shown in dotted lines.

Associations between covariates and the risk of reinfection remained broadly similar in sensitivity analyses that further adjusted for background infection prevalence, with only small differences for the effects of age and region (**Figure S5, Figure 4, Table S4A**). A 1% higher background prevalence was associated with a 20%, 23%, 33%, and 27% higher risk of reinfection in the Omicron BA.1, BA.2, BA.4/5, and BQ.1/CH.1.1/XBB.1.5 waves (**Table S4A**). Considering participants as being ‘at risk’ from the date of their first negative PCR test in the study following each infection rather than 120 days after their previous infection (see **Methods**) generated similar results; being <120 days from the previous infection was consistently associated with a much lower risk of reinfection in different waves (**Table S4B, Figure S8, Figure S9**). The associations with sex, ethnicity, and working in healthcare largely disappeared when only including infections defined by positive test results from the study (so not influenced by test seeking behaviour), but protection from previous infection remain similar (**Table S4C**).

## Discussion

Here, we have quantified the high reinfection rates associated with multiple Omicron waves driven by the BA.1, BA.2, BA.4/5, and the most recent BQ.1/CH.1.1/XBB.1.5 variants. Previous studies found the reinfection rate with pre-Omicron variants was less than 1%^23,24^, while others have described reinfection rates with Omicron BA.1/BA.2 of 6%-15%^17,25,26^. In our population, the percentage of those previously infected who became reinfected was also around 10-11% at the end of the BA.1 and BA.2 waves, but reached 14-16% at the end of the BA.4/5 and BQ.1/CH.1.1/XBB.1.5 waves, likely reflecting both the longer duration of these waves and viral immune evasion by most recent variants (**Figure 2)^27,28^**. We also found that around 1 in 70 participants were identified as having been infected three times, and 1 in 1500 participants four or five times over the three year study period, with most fourth or fifth reinfections in the Omicron BA.4/5 and BQ.1/CH.1.1/XBB.1.5 waves.

Re-infections were somewhat less likely to be symptomatic, with 40-50% decreased risk of reporting classic symptoms in reinfections versus first infections; however, one limitation is that symptoms were self-reported and inevitably contain an element of subjectivity as well as confounding with other infections. Reinfections also had higher Ct values, including after adjustment for infection variants, suggesting that reinfections were generally less severe, regardless of variants, as Ct values are inversely associated with viral load, which has been associated with the severity and outcomes of SARS-CoV-2 infections^29^. This is consistent with a previous systematic review reporting reinfections were more likely to cause mild illness and were associated with a lower risk of hospitalisation and death compared with the first infections^30^. However, SARS-CoV-2 reinfection may still pose an additional risk of death and hospitalization in people aged >60 years, which may be related to reduced immunity and comorbidities in this group^31^. Therefore, prevention of SARS-CoV-2 reinfection remains particularly important for the older and other high-risk populations.

Using flexible parametric survival models, we estimated the risk of reinfection in multiple Omicron infection waves, and specifically examined the protection afforded by previous infection and vaccination. Across all four Omicron waves, we found that protection against reinfection was greatest following previous infection with the most recent vs earlier Omicron variants, and with Omicron vs pre-Omicron infections, even at the same time from previous infection (**Figure 2**). Because variants emerge and calendar time passes together, it is potentially challenging to distinguish whether effects are from changes in variant per se or the time from the most recent infection. However, where the time since the most recent infection overlapped, reinfection risks associated with different variants showed risk was still generally lower with a more recent variant than an earlier variant at any given time since infection. Given SARS-CoV-2 has a great ability to evolve and accumulate genetic diversity, these data also indicate that viral evolution is likely as great a driver of reinfection as the waning of host immunity.

We found significant waning over time of protection against BA.4/5 reinfection following BA.1, BA.2 infections, and against BQ.1/CH.1.1/XBB.1.5 reinfection following BA.2 and BA.4/5 infections, consistent with other reports of protection against Omicron reinfection decreasing over time from previous Omicron infections^32^. We did not find any evidence of waning of protection against a new Omicron reinfection if the most recent infection was with a Pre-Alpha, Alpha, or Delta variant, excepting only previous Delta infections against BA.2 reinfections. However, many of these Pre-Alpha/Alpha/Delta previous infections were >360 days previously, particularly for later Omicron waves. Our findings are consistent with waning of protection from previous infection with any variant plateauing after a year, and almost complete immune escape with Omicron BA.1. The risk of BQ.1/CH.1.1/XBB.1.5 reinfection was similar or even slightly lower 540 days after a most recent Pre-Alpha or Alpha infection than that after a most recent Delta or BA.1 infection, with similar trends for BA.4/5 reinfections. This may be explained by a ‘healthy survivor effect’, as participants with a Pre-Alpha/Alpha infection who remained free from being reinfected in the early three Omicron waves may have better immunity in general and/or have specific behaviours that confer protection (e.g. continued shielding).

We found that the risk of reinfection increased over time from the most recent vaccination in BA.1, BA.4/5, and BQ.1/CH.1.1/XBB.1.5 waves. Reduced risk of reinfection after vaccination was reported previously for Alpha, Delta, and BA.1 variants^26,33,34^, suggesting that among people with previous infection, vaccination could provide additional protection against reinfection, and this protection decreased over time. This is consistent with our results that those receiving a vaccination <6 months previously had a lower risk of reinfection than those >6 months, indicating that people previously infected could still benefit from a more recent vaccination. The lack of evidence for an association in the BA.2 wave could represent lower power (as effect estimates numerically favoured benefit), a true lack of additional benefit from vaccination with this variant, or unmeasured confounding among those clinically vulnerable individuals who should have recently received a fourth dose during the BA.2 wave and would be at a higher risk of reinfection.

We also found that the risk of reinfection was consistently higher in females, those of white ethnicity, and healthcare workers across different waves, but a sensitivity analysis restricting definitions of infection outcomes and exposures to be based only on tests done within the study suggested this could mostly be explained by a more frequent test seeking behaviour in these groups. Reporting classic symptoms in the most recent previous infection was associated with a lower risk of BA.1 and BA.2 reinfections, but not BA.4/5 and BQ.1/CH.1.1/XBB.1.5 reinfections. The risk of reinfection was much lower in Northern Ireland, Scotland, and Wales than in England during BA.1 and BA.2 waves but was similar during the BA.4/5 wave across regions and higher in Northern Ireland and Wales than England during the most recent BQ.1/CH.1.1/XBB.1.5 wave. These effects were attenuated adjusting for background prevalence, suggesting that they were driven by increased transmission risk overall, rather than being specific to reinfections.

We also found a non-linear U-shaped effect on the risk of reinfection associated with age and Ct values at the most recent previous infection. The risk was higher among those aged 30-45 years, consistent with previous studies on Delta variant^35^ and may be explained by working-age adults having greater exposure to social and inter-generational interactions and thus having a higher transmission rate. The lower risk of reinfection in older participants may be due to both lower social interactions and vaccination policy, as they were the first to receive a third booster vaccination at the end of 2021 and those 75 years and older were also given further booster vaccinations in the first quarter of 2022. The reinfection risk was higher in participants with either a low or high Ct value in their most recent previous infection. For those with a very high measured Ct value in their first infection, potentially a relatively less severe initial infection may not generate an effective immune response, leaving these participants more likely to be reinfected. In contrast for those with a very low Ct value, the higher viral load in the first infection may indicate a lack of initial viral control that may also be associated with susceptibility to a subsequent infection.

Study strengths include the very large size and unbiased sampling frame of the study, allowing us to unambiguously decompose the relative contribution of different types of previous infection, and time from these previous infections, on reinfection risk. We also used calendar time as our underlying time-scale in time-to-event models, to allow for the fact that background infection pressure changes substantially, even over the course of a wave (**Figure S4**), and could lead to unmeasured confounding if not adjusted for sufficiently flexibly. We also carefully considered other biases, including immortal time bias. Study limitations include the fact that our primary analysis chose not to estimate the reinfection risk within 120 days from the most recent previous infection because of challenges identifying short reinfections (requiring sequencing and/or reliable S-gene positivity data, available only for tests with Ct<30). Our main analyses therefore excluded a number of short reinfections, predominantly with Omicron BA.1 and BA.2. However, results remained very similar in sensitivity analyses including participants from their first negative study PCR test, rather than from 120 days after their previous infection, in order to incorporate shorter reinfections. Although the study design was to assess participants every 28-42 days regardless of symptomatology, and most intervals between assessments were <45 days, it is inevitable that we still missed some infections. However, we used positive PCR test results from the study, positive test results from England and Wales’s National Testing programme, and self-reported positive tests to define infection episodes and reduce misclassification of both exposure and outcome. However, this means that test seeking behaviour could have an effect on estimates, although this should be minimised by including study results. Ct values were missing from 37.1% of infections, and Ct values could also be influenced by the timing of infection detection in the study, since tests were performed regardless of symptom. We used pre-specified rules to define reinfections based on sequencing data, S-gene presence/absence, Ct values and calendar time (see Methods), but these rules were not perfect, especially for those shorter reinfections, and sequencing could only be attempted on a subset of infections with low Ct. We used calendar time to define the variant of previous infections for those without sequencing data and with only one or no gene-positive, which may be subject to misclassification bias. Symptoms were underreported before the study data collection became digital in June 2022, so the associations between symptoms in reinfections vs first infections could be underestimated. We did not have data on hospitalisation and death, thus could not assess disease severity as outcomes. Lastly, the risk of reinfection assessed as an outcome in the main analyses inevitably includes both the risk of being exposed to the virus and the risk of being reinfected given exposure. Given the background infection rate was associated with the risk of virus exposure, a sensitivity analysis adjusted for infection prevalence by calendar time, age, and region. As expected, a higher prevalence was associated with a higher risk of reinfection, but effects from other covariates, especially time from the most recent infection, remained very similar.

In conclusion, the risk of reinfection in the Omicron waves is high, and is likely driven as much by viral evolution as waning immunity, but reinfections are generally less severe than the first infections. A previous Omicron infection provides higher protection than a previous pre-Omicron infection against new Omicron reinfections, but this protection decreases over time. Additional protection from vaccination also decreases over time. Given the waning immunity from previous infection and vaccination, and the stronger immune evasion from most recent Omicron variants, reinfection risk remains high, and ongoing evaluation of hybrid immunity against emerging dominant variants remains important to pandemic control and vaccination policy. Further public health measures may be needed to help protect vulnerable people from reinfection.

## Methods

### Population and setting

The ONS COVID-19 Infection Survey (CIS) is a large household survey with longitudinal follow-up (ISRCTN21086382; https://www.ndm.ox.ac.uk/covid-19/covid-19-infection-survey/protocol-and-information-sheets). Private households were randomly selected from address lists and previous surveys on a continuous basis for enrolment from 26 April 2020 through 31 January 2022 (when new recruitment was paused, although follow-up continued until 13 March 2023 when study assessments were paused). Following verbal agreement to participate, a study worker visited each selected household to take written informed consent for individuals aged 2y and over. For those aged 2-15y, consent was provided by their parents or carers; those 10–15y also provided written assent. At the first visit, participants were asked for consent for optional follow-up assessments every week for the next month and then monthly subsequently. The study received ethical approval from the South Central Berkshire B Research Ethics Committee (20/SC/0195).

At each assessment, participants were asked about demographics, behaviours, work, and vaccination status. Combined nose and throat swabs were taken from all consenting household members for SARS-CoV-2 PCR testing. Blood samples were taken monthly for antibody testing from participants aged 16y and over in a randomly selected 10-20% of households. Household members of participants who tested positive on a nose and throat swab were also invited to provide blood monthly for follow-up assessments. From April 2021, additional participants were invited to provide blood samples monthly to assess vaccine responses, based on a combination of random selection and prioritization of those in the study for the longest period (independent of swab test results). Details on the sampling design are provided elsewhere^36^. From 14 July 2022, study worker visits were discontinued and participants could opt-in to continuing to complete questionnaires online or by telephone, returning test kits by post or courier^37–39^. Data collection was officially paused on 13 March 2023.

### Vaccination data

Self-reported vaccination data were obtained from participants at assessments, including vaccination type, number of doses, and vaccination dates. Data from participants in England and Wales were also linked to administrative records from the National Immunisation Management Service (NIMS) in England and equivalent in Wales. We used records from administrative data sources where available and otherwise from the study, since linkage was periodic and administrative data sources do not contain information about vaccinations received abroad or in Northern Ireland and Scotland.

### Laboratory testing

Combined nose and throat swabs from the CIS were analysed at the UK’s national Lighthouse Laboratories at Milton Keynes and Glasgow using identical methodology. PCR for three SARS-CoV-2 genes (N protein, S protein, and ORF1ab) was performed using the Thermo Fisher TaqPath RT-PCR COVID-19 kit, and analysed using UgenTec FastFinder 3.300.5, with an assay-specific algorithm and decision mechanism that allows conversion of amplification assay raw data from the ABI 7500 Fast into test results with minimal manual intervention. Positive samples are defined as having at least a single N and/or ORF1ab gene detected, and PCR traces exhibited an appropriate morphology. The S gene alone is not considered to be positive^36^. These test results had both gene positivity and cycle threshold (Ct) values available. During periods of high sample returns, a small number of CIS swabs were tested at two other laboratories, one using an endpoint PCR test (no Ct or gene positivity data available). For participants in England and Wales, we also included positive swab test results (SARS-CoV-2 PCR and lateral flow device tests) linked from national clinical/hospital-based testing (also including health and care workers) and community testing programmes^40^. A substantial proportion of these additional tests were performed at the Lighthouse Laboratories using the same test as used in CIS: we used gene positivity and Ct values from these laboratories but not other laboratories in the national testing programme. As these linked positive test results were not available for participants from Scotland and Northern Ireland, we also included self-reported positive swab tests.

### SARS-CoV-2 reinfection definition

We grouped repeated positive tests from the sources above into infection episodes to reflect the fact that some individuals test positive on PCR for extended periods of time, incorporating information from genetic sequencing, S-gene presence/absence, and cycle threshold (Ct) values, together with negative PCR test results from CIS only. Using criteria developed through expert consensus, careful inspection and analysis of CIS data alone, and considering definitions of reinfection used elsewhere^15,31,41,42^, we defined the start of a new infection episode as any of the following: 1) a new swab positive occurring >120 days after an index positive with the preceding test being negative based on analyses of vaccine effectiveness against Delta and Alpha variants which showed that definitions based on shorter periods of time and/or without a previous negative in these earlier calendar periods erroneously included those testing PCR-positive for long periods of time^43^, or 2) >90 days with the preceding two consecutive tests being negative (one negative after 20 December 2021 when Omicron variants dominated given higher re-infection rates with Omicron^15,17^), or 3) >60 days with the three preceding consecutive tests being negative, or 4) after 4 preceding consecutive negative test results at any time.

We then split these infection episodes if they had grouped together positive tests containing multiple sequences from different genetic lineages (e.g. BA.5 and BA.2), or had incompatible S-gene target positivity consistent with co-circulating variants with Ct<30 (e.g. S-gene positive and S-gene negative, both with Ct<30 during periods when BA.1 and BA.2 were co-circulating), or had large decreases in Ct or low Ct long after the first positive within an episode (both indicative of a new infection rather than ongoing PCR positivity). We also split infection episodes where a new lateral flow device positive was recorded 19 days or more after the first positive in an infection episode, since this again indicates high viral load and actively replicating virus, more likely associated with a new infection. We added a small number of additional infections identified by a positive S-antibody test occurring before vaccination and at least 90 days before the first swab-positive episode.

### SARS-CoV-2 reinfection variants, Ct values and symptoms

The variant associated with each infection episode was determined by whole genome sequencing where this was available, otherwise as Pre-Alpha/Delta/Omicron BA.2-compatible if the S-gene was detected (with N/ORF1ab/both), or as Alpha/Omicron BA.1/Omicron BA.4/5-compatible if positive at least once for ORF1ab+N (but not for the S gene, S-gene target failure, SGTF), using the dates when these variants were dominant in the UK, i.e. accounted for >50% of infections. For those without sequencing data and with only one gene-positive (N-only/ORF1ab-only) or no gene positivity/Ct data available, we assigned them to each variant type based on the national dominant circulating variant in each surveillance week (>50% of positive tests in the study): Pre-Alpha (before 06 December 2020), Alpha (07 December 2020 to 16 May 2021), Delta (17 May 2021 to 12 December 2021), Omicron BA.1 (13 December 2021 to 20 February 2022), Omicron BA.2 (21 February 2022 to 5 June 2022), and Omicron BA.4/5 infections (6 June 2022 to 6 November 2022). After 7 November 2022, sequencing data in the UK showed a mixture of different sub-lineages including the BQ.1 variant (a sub-lineage of BA.5), CH.1.1 variant (a sub-lineage of BA.2.75), and XBB variant (a recombinant lineage derived from two BA.2 sub-lineages), so this wave was denoted ‘BQ.1/CH.1.1/XBB.1.5’.

In addition to defining variants associated with each infection as exposures, we used a stricter rule to define variant waves as outcomes to allow separate reinfection survival models to be fitted for new infections and reduce misclassification bias for this outcome, based on time periods where a given variant accounted for ≥85% of infections: Omicron BA.1 (27 December 2021 to 6 February 2022), Omicron BA.2 (14 March 2022 to 22 May 2022), Omicron BA.4/5 (27 June 2022 to 6 November 2022), and Omicron BQ.1/CH.1.1/XBB.1.5 subvariants (7 November 2022 to 13 March 2023).

The Ct value associated with each infection was the minimum observed across all positive tests within each episode performed using the same test at the Lighthouse laboratories, taking the mean Ct value across all detected targets per test (and then the minimum across tests).

Associated symptoms were defined as all symptoms from the list below reported by participants within [0,35] days of the first positive test in each infection episode^44^, including symptoms reported in the last 7 days at assessments and symptoms reported at dates when participants reported they had had COVID. These symptoms were classified as ‘classic’ (any of cough, fever, loss of taste/smell) or ‘other’ (any of another 8 symptoms solicited consistently since the start of the study (myalgia, fatigue/weakness, sore throat, shortness of breath, headache, diarrhoea, nausea, abdominal pain))^45^. If no symptoms were reported within CIS, we also included any self-reported symptoms available from routine national testing as “other” as specific symptoms were not available from these data sources.

### Statistical analysis

We included participants ≥18y at their first infection episode who had at least one infection episode from 28 February 2020 (first positive swab test in linked or self-reported data) to 13 March 2023. Data from all eligible participants were included, but age was truncated at 85 years for those >85 years to reduce the influence of outliers (0.7% of all participants).

We first investigated how Ct values (a surrogate for viral load and potentially infection severity; lower Ct values indicate higher viral loads^46^) varied by infection episode (initial infection, first reinfection, second reinfection) using a robust linear regression model, adjusting for the variant of the current infection, age, sex, ethnicity, reporting working in patient-facing healthcare, report having a long-term health condition, deprivation percentile, self-reported symptoms (classic, other) in the current infection episode, number of positive tests in an infection episode, and time from the most recent vaccination (no vaccination, <14 days, 14-90 days, 90-180 days, and >180 days). Pairwise interactions between the infection episode, variant, and time from the most recent vaccination were included to examine the relationships between these different factors and Ct values as these were highly significant. A separate robust linear regression model estimated associations between the Ct value of reinfections (second and third infections) and Ct value and self-reported symptoms in the most recent previous infection. Other covariates remained the same. Ct values in the previous infection were fitted using restricted cubic splines with 3 knots to account for nonlinearity.

We then examined how the percentage of infections where participants reported symptoms (no symptom, classic symptoms, other symptoms) varied by infection episode (initial infection, first reinfection, second reinfection) using a multinomial regression model, adjusting for the same covariates as above: variant of the current infection episode, age, sex, ethnicity, reporting working in patient-facing healthcare, report having a long-term health condition, deprivation percentile, Ct values in the current infection episode, and time from the most recent vaccination (no vaccination, <14 days, 14-90 days, 90-180 days, and >180 days). Interactions were not included because there was no evidence suggesting that including the interaction terms improved model fit.

We used time-to-event survival models to estimate the risk of reinfection, treating calendar date as the underlying time scale and using flexible parametric survival models (stpm2)^47,48^ to reflect the substantial changes in background infection rate with each wave, modelling the baseline log-cumulative hazard using B-splines. The optimal number of knots for the splines was selected based on minimizing the Akaike information criterion (AIC). Separate survival models were built for reinfections that happened during each variant wave (defined above), using the date of start of the wave as time 0. Counting participants as being “at risk” from their previous index positive date could cause ‘immortal time bias’^49^ because reinfections could not happen until the participant stopped testing positive repeatedly. Whilst in theory subsequent positive tests could be assigned to a new infection episode on the basis of sequencing, S-gene presence/absence and/or Ct values as described above, as sequencing was only performed and S-gene presence/absence can only be reasonably reliably detect in samples with low Ct, potential for this immortal time bias remains. Therefore, we defined an ‘at-risk date’ following each infection as the first date a participant could have been counted as having reinfection had they tested positive based only on the sliding scale of time and number of previous negative tests. Each participant entered the risk set at the later of the start of the wave (time 0) or the date they were first at-risk of reinfection using a late-entry method. They were censored at the last date of the wave, or the last known date of their study assessments, whichever was earlier. Thus analyses included as both exposures and outcomes previous infections only identified through non-study tests, but not infections identified after study participation stopped. 67% of participants with any infection were recruited before 7 December 2020, the start of the Alpha wave.

We then excluded time at risk <120 days from previous infection to minimise immortal time bias from varying durations of observed PCR positivity, thus ensuring that risk sets were not very small and unrepresentative shortly after previous infection. This excluded a number of observed reinfections from each wave (603 (17%) Omicron BA.1, 1,194 (21%) Omicron BA.2, 720 (5%) Omicron BA.4/5, 348 (2%) Omicron BQ.1/CH.1.1/XBB.1.5). For sensitivity analyses, we counted participants as being ‘at risk’ from the date of their first negative study PCR test (i.e. not excluding time at risk <120 days from a previous infection and thus including shorter reinfections).

We used a multivariable model to examine associations between risk of reinfection and continuous age (16-85 years), sex, ethnicity (white vs. non-white due to small numbers), reporting having a long-term health condition, reporting working in patient-facing healthcare, deprivation percentile, geographical region (England, Scotland, Wales, Northern Ireland), Ct values in the most recent previous infection, symptoms in the most recent previous infection (classic and other symptoms), had any Ct<30 or lateral flow device (LFD) positive in previous infections, and the number of previous infections. Age and Ct values were fitted using restricted cubic splines with 3 knots to account for non-linear effects. To examine waning immunity from natural infection or vaccination, we further added two time-varying covariates: time from previous infection (split into 120-180, 180-240, 240-300, 300-360, 360-420, 420-480, 480-540, 540-600, 600-660, 660-720, >720 days), and time from most recent vaccination (no vaccination, 14-90 days, 90-180 days, >180 days after the most recent vaccination). Time from previous infection was modelled categorically and as a trend across these categories in separate models.

For sensitivity analyses, we further adjusted for infection prevalence in the survival models to account for the influence from the background infection pressure. In this way, we could try to separate the risk of being exposed to the virus from those being reinfected given exposure. A generalised additive model (GAM) was built using all study swab PCR test results as the denominator and positive study PCR results as the outcome. Calendar time and age were included using a tensor product spline which was allowed to vary by region/country. Predicted values from the GAM model were used to represent the prevalence varied by calendar time, age, and region, which were then joined to the dataset and included as a covariate in the survival models.

All analyses were performed in R 4.0 using the following packages: tidyverse (version 1.3.1), MASS (version 7.3-53), arsenal (version 3.4.0), cowplot (version 1.1.1), survival (version 3.2-7), and rstpm2 (version 1.5.2).

## Data Availability

De-identified study data are available for access by accredited researchers in the ONS Secure Research Service (SRS) for accredited research purposes under part 5, chapter 5 of the Digital Economy Act 2017. Individuals can apply to be an accredited researcher using the short form on https://researchaccreditationservice.ons.gov.uk/ons/ONS_registration.ofml. Accreditation requires completion of a short free course on accessing the SRS. To request access to data in the SRS, researchers must submit a research project application for accreditation in the Research Accreditation Service (RAS). Research project applications are considered by the project team and the Research Accreditation Panel (RAP) established by the UK Statistics Authority at regular meetings. Project application example guidance and an exemplar of a research project application are available. A complete record of accredited researchers and their projects is published on the UK Statistics Authority website to ensure transparency of access to research data. For further information about accreditation, contact or visit the SRS website.

## Acknowledgements

We are grateful for the support of all COVID-19 Infection Survey participants.

This study is funded by the UK Health Security Agency and the Department of Health and Social Care with in-kind support from the Welsh Government, the Department of Health on behalf of the Northern Ireland Government and the Scottish Government. JW is supported by University of Oxford and the China Scholarship Council. ASW, TEAP, NS, KBP are supported by the National Institute for Health Research Health Protection Research Unit (NIHR HPRU) in Healthcare Associated Infections and Antimicrobial Resistance at the University of Oxford in partnership with the UK Health Security Agency (UK HSA) (NIHR200915). ASW and TEAP are also supported by the NIHR Oxford Biomedical Research Centre. KBP is also supported by the Huo Family Foundation. ASW is also supported by core support from the Medical Research Council UK to the MRC Clinical Trials Unit [MC_UU_12023/22] and is an NIHR Senior Investigator. PCM is funded by Wellcome (intermediate fellowship, grant ref 110110/Z/15/Z) and holds an NIHR Oxford BRC Senior Fellowship award. DWE is supported by a Robertson Fellowship and an NIHR Oxford BRC Senior Fellowship. NS is an Oxford Martin Fellow and holds an NIHR Oxford BRC Senior Fellowship. The views expressed are those of the authors and not necessarily those of the National Health Service, NIHR, Department of Health, or UKHSA.

## The COVID-19 Infection Survey team

John N Newton^11^, John I Bell^12^, Jeremy Farrar^13^, Jaison Kolenchery^1,5^, Brian D. Marsden^1,14^, Sarah Hoosdally^1^, E Yvonne Jones^1^, David I Stuart^1^, Derrick W. Crook^1,5,6,7^, Tina Thomas^9^, Daniel Ayoubkhani^9^, Russell Black^9^, Antonio Felton^9^, Megan Crees^9^, Joel Jones^9^, Lina Lloyd^9^, Esther Sutherland^9^, Emma Pritchard^1^, Karina-Doris Vihta^1^, George Doherty^1^, James Kavanagh^1^, Kevin K. Chau^1^, Stephanie B. Hatch^1^, Daniel Ebner^1^, Lucas Martins Ferreira^1^, Thomas Christott^1^, Wanwisa Dejnirattisai^1^, Juthathip Mongkolsapaya^1^, Sarah Cameron^1^, Phoebe Tamblin-Hopper^1^, Magda Wolna^1^, Rachael Brown^1^, Richard Cornall^1^, Gavin Screaton^1^, Katrina Lythgoe^2^, David Bonsall^2^, Tanya Golubchik^2^, Helen Fryer^2^, Stuart Cox^15^, Kevin Paddon^15^, Tim James^15^, Thomas House^16^, Julie Robotham^17^, Paul Birrell^17^, Helena Jordan^18^, Tim Sheppard^18^, Graham Athey^18^, Dan Moody^18^, Leigh Curry^18^, Pamela Brereton^18^, Ian Jarvis^19^, Anna Godsmark^19^, George Morris^19^, Bobby Mallick^19^, Phil Eeles^19^, Jodie Hay^20^, Harper VanSteenhouse^20^, Jessica Lee^21^, Sean White^22^, Tim Evans^22^, Lisa Bloemberg^22^, Katie Allison^23^, Anouska Pandya^23^, Sophie Davis^23^, David I Conway^24^, Margaret MacLeod^24^, Chris Cunningham^24^

^11^European Centre for Environment and Human Health, University of Exeter, Truro, UK

^12^Office of the Regius Professor of Medicine, University of Oxford, Oxford, UK

^13^Wellcome Trust, London, UK

^14^Nuffield Department of Orthopaedics, Rheumatology and Musculoskeletal Sciences, University of Oxford, Oxford, UK

^15^Oxford University Hospitals NHS Foundation Trust, Oxford, UK

^16^University of Manchester, Manchester, UK

^17^Health Improvement Directorate, Public Health England, London, UK

^18^IQVIA, London, UK

^19^National Biocentre, Milton Keynes, UK.

^20^Glasgow Lighthouse Laboratory, London, UK

^21^Department of Health and Social Care, London, UK

^22^Welsh Government, Cardiff, UK

^23^Scottish Government, Edinburgh, UK

^24^Public Health Scotland, Edinburgh, UK

## Author Contributions

The study was designed and planned by ASW, ID and KBP, and is being conducted by ASW, TK, OG, RS, NT, TEAP, PCM, NS, DWE, and the COVID-19 Infection Survey Team. This specific analysis was designed by JW, DWE, ASW, and KBP. JW contributed to the statistical analysis of the survey data. JW, DWE, KBP and ASW drafted the manuscript and all authors contributed to interpretation of the data and results and revised the manuscript. DWE, KBP, and ASW contributed equally. All authors approved the final version of the manuscript.

