## supplementary for "Risk of SARS-CoV-2 reinfection during multiple Omicron variant waves in the UK general population"

### Supplementary results

|  |  |  |  | First infection | | | Second infection | | | Third infection | | | Fourth infection | | | Fifth infection | | |
| --- | --- | --- | --- | --- | --- | --- | --- | --- | --- | --- | --- | --- | --- | --- | --- | --- | --- | --- |
|  | **Age range at first infection** | **Sex** | **Ethnicity** | Variant | Symptom | Ct value/LFD positive | Variant | Symptom | Ct value/LFD positive | Variant | Symptom | Ct value/LFD positive | Variant | Symptom | Ct value/LFD positive | Variant | Symptom | Ct value/LFD positive |
| 1 | **46-50** | **Female** | **White** | Pre-Alpha | Classic | 32 | Delta | Classic | Not positive | Delta | Other | Not positive | BA.2 | Classic | Positive | BA.4/5 | No | Positive |
| 2 | **46-50** | **Female** | **White** | AY.4* | Classic | Not positive | BA.1 | Other | Not positive | BA.2 | Classic | Positive | BA.4/5 | Classic | Not positive | BA.5.1* | Classic | Positive |
| 3 | **36-40** | **Female** | **White** | Delta | Other | Positive | BA.2 | No | Positive | BA.4/5 | No | Positive | BA.4/5 | No | Positive | CA.7* | Classic | Not positive |
| 4 | **21-25** | **Female** | **White** | Delta | Other | Not positive | BA.1 | No | Positive | BA.2 | Classic | Not positive | BF.27* | No | Positive | BQ.1/CH.1.1/XBB.1.5 | Classic | Not positive |
| 5 | **36-40** | **Female** | **White** | Pre-Alpha | Classic | 29 | Delta | No | 28 | Delta | Other | Positive | BA.1* | Other | 18 | BA.4/5 | No | Positive |
| 6 | **36-40** | **Female** | **White** | Pre-Alpha | Classic | Not positive | BA.1.1* | Classic | 18 | BA.2 | Classic | Positive | BQ.1/CH.1.1/XBB.1.5 | Classic | Positive | BE.9* | Classic | 16 |
| 7 | **41-45** | **Female** | **White** | Delta | Other | Not positive | BA.1 | Other | 21 | BA.2 | Other | Not positive | BA.4/5 | Classic | Not positive | BQ.1.1* | Classic | 28 |

**Table S1. Characteristics of infections in 7 participants who had five infections.** We used pangolin sequence assignment if available (marked as *), otherwise defined the infection variant based on calendar time (see Methods). The Ct value associated with each infection was the minimum observed across all positive tests within each infection (see Methods). If no Ct was recorded, we classified the infection by whether there were any lateral flow device (LFD) positive results from the national testing programmes in the infection. Symptoms were classified as ‘classic’ (any of cough, fever, loss of taste/smell) or ‘other’ (myalgia, fatigue/weakness, sore throat, shortness of breath, headache, diarrhoea, nausea, abdominal pain). If no symptoms were reported within CIS, we included any self-reported symptoms available from national testing programmes as “other” as specific symptoms were not available.

|  | Model including first, second, and third infections | | | | Model including only second and third infections | | | |
| --- | --- | --- | --- | --- | --- | --- | --- | --- |
|  | Difference in Ct value in current infection | 95%CI | | p value | Difference in Ct value in current infection | 95%CI | | p value |
| Age (per 10-year older) | -0.037 | -0.056 | -0.017 | **<0.001** | 0.090 | 0.027 | 0.152 | **0.005** |
| Sex (Male vs Female) | -0.490 | -0.547 | -0.432 | **<0.001** | -0.483 | -0.649 | -0.317 | **<0.001** |
| Ethnicity (Non-white vs White) | -0.079 | -0.192 | 0.034 | 0.2 | -0.013 | -0.319 | 0.293 | 0.8 |
| Working in healthcare (Yes vs No) | -0.217 | -0.347 | -0.088 | **0.001** | -0.201 | -0.559 | 0.156 | 0.3 |
| Having a long-term health condition (Yes vs No) | 0.050 | -0.017 | 0.118 | 0.1 | -0.183 | -0.385 | 0.018 | 0.07 |
| Deprivation percentile (per 10 percentile higher) | 0.029 | 0.018 | 0.040 | **<0.001** | 0.000 | -0.030 | 0.030 | 0.9 |
| Reported symptoms in this infection (Classic vs No) | -3.918 | -3.990 | -3.846 | **<0.001** | -2.797 | -2.992 | -2.603 | **<0.001** |
| Reported symptoms in this infection (Other vs No) | -3.369 | -3.460 | -3.278 | **<0.001** | -1.930 | -2.185 | -1.675 | **<0.001** |
| Reported symptoms in previous infection (Classic vs No) |  |  |  |  | 0.210 | 0.001 | 0.419 | **0.04** |
| Reported symptoms in previous infection (Other vs No) |  |  |  |  | 0.151 | -0.086 | 0.387 | 0.2 |
| Number of positive tests in the infection (per one more) | -0.125 | -0.155 | -0.095 | **<0.001** | 0.377 | 0.232 | 0.522 | **<0.001** |
| Variant (Pre-Alpha vs Omicron BA.1) | 4.730 | 2.584 | 6.877 | **<0.001** | 5.895 | 0.346 | 11.444 | **0.04** |
| Variant (Alpha vs Omicron BA.1) | 1.581 | -2.207 | 5.368 | **0.4** | 5.656 | 4.023 | 7.289 | **<0.001** |
| Variant (Delta vs Omicron BA.1) | -0.732 | -0.989 | -0.475 | **<0.001** | 0.016 | -1.315 | 1.347 | 0.8 |
| Variant (Omicron BA.2 vs Omicron BA.1) | 1.783 | 1.554 | 2.011 | **<0.001** | 2.595 | 1.950 | 3.240 | **<0.001** |
| Variant (Omicron BA.4/5 vs Omicron BA.1) | 2.425 | 2.227 | 2.624 | **<0.001** | 2.386 | 1.840 | 2.932 | **<0.001** |
| Variant (Omicron BQ.1/CH.1.1/XBB.1.5 vs Omicron BA.1) | 3.407 | 3.169 | 3.645 | **<0.001** | 3.344 | 2.792 | 3.897 | **<0.001** |
| Time from most recent vaccination (No vaccination vs >180 days) | -0.086 | -0.524 | 0.353 | 0.7 | -0.129 | -1.125 | 0.867 | 0.8 |
| Time from most recent vaccination (14-90 days vs >180 days) | 0.227 | 0.035 | 0.420 | **0.02** | 1.089 | 0.493 | 1.686 | **<0.001** |
| Time from most recent vaccination (90-180 days vs >180 days) | 0.452 | 0.247 | 0.658 | **<0.001** | 1.076 | 0.415 | 1.738 | **0.001** |
| Infection (2 vs 1) | 0.834 | 0.564 | 1.104 | **<0.001** |  |  |  |  |
| Infection (3 vs 1) | 2.432 | 1.021 | 3.842 | **<0.001** | 0.329 | 0.007 | 0.650 | **0.04** |
| Variant: Time from most recent vaccination (interaction) |  |  |  | **<0.001** |  |  |  | **<0.001** |
| Pre-Alpha:No vaccination | -2.221 | -4.404 | -0.039 | **0.04** | -1.597 | -7.710 | 4.516 | 0.6 |
| Alpha:No vaccination | 0.705 | -3.103 | 4.513 | 0.7 | -1.556 | -3.777 | 0.665 | 0.2 |
| Delta:No vaccination | 0.288 | -0.281 | 0.856 | 0.3 | 1.077 | -1.183 | 3.338 | 0.4 |
| Omicron BA.2:No vaccination | -0.697 | -1.373 | -0.020 | **0.04** | -1.579 | -3.033 | -0.125 | **0.03** |
| Omicron BA.4/5:No vaccination | 0.574 | -0.255 | 1.402 | 0.2 | 0.752 | -0.739 | 2.243 | 0.3 |
| Omicron BQ.1/CH.1.1/XBB.1.5:No vaccination | 0.005 | -0.908 | 0.917 | 0.9 | -0.048 | -1.590 | 1.494 | 0.9 |
| Alpha:14-90 days | 2.193 | -1.614 | 6.001 | 0.3 |  |  |  |  |
| Delta:14-90 days | 0.249 | -0.055 | 0.552 | 0.1 | 1.990 | 0.379 | 3.600 | **0.02** |
| Omicron BA.2:14-90 days | -0.972 | -1.251 | -0.694 | **<0.001** | -1.812 | -2.653 | -0.970 | **<0.001** |
| Omicron BA.4/5:14-90 days | -0.168 | -0.478 | 0.143 | 0.3 | -0.399 | -1.280 | 0.482 | **0.4** |
| Omicron BQ.1/CH.1.1/XBB.1.5:14-90 days | -0.301 | -0.583 | -0.019 | **0.04** | -1.060 | -1.745 | -0.376 | **0.002** |
| Delta:90-180 days | -0.430 | -0.728 | -0.132 | **0.005** | -1.315 | -2.884 | 0.253 | 0.1 |
| Omicron BA.2:90-180 days | -1.010 | -1.268 | -0.751 | **<0.001** | -1.893 | -2.700 | -1.085 | **<0.001** |
| Omicron BA.4/5:90-180 days | -0.970 | -1.274 | -0.666 | **<0.001** | -1.604 | -2.470 | -0.738 | **<0.001** |
| Omicron BQ.1/CH.1.1/XBB.1.5:90-180 days | -0.714 | -1.011 | -0.417 | **<0.001** | -1.317 | -2.067 | -0.568 | **<0.001** |
| Variant:Infection (interaction) |  |  |  | **<0.001** |  |  |  |  |
| Pre-Alpha:infection 2 | 1.988 | 0.029 | 3.947 | **0.04** |  |  |  |  |
| Alpha:infection 2 | 1.810 | 0.808 | 2.812 | **<0.001** |  |  |  |  |
| Delta:infection 2 | 0.808 | 0.321 | 1.295 | **0.001** |  |  |  |  |
| Omicron BA.2:infection 2 | 0.014 | -0.269 | 0.297 | 0.9 |  |  |  |  |
| Omicron BA.4/5:infection 2 | -0.289 | -0.588 | 0.010 | 0.06 |  |  |  |  |
| Omicron BQ.1/CH.1.1/XBB.1.5:infection 2 | -0.424 | -0.693 | -0.155 | **0.002** |  |  |  |  |
| Delta:infection 3 | -2.112 | -6.833 | 2.609 | **0.4** |  |  |  |  |
| Omicron BA.2:infection 3 | -1.451 | -3.038 | 0.135 | 0.07 |  |  |  |  |
| Omicron BA.4/5:infection 3 | -1.576 | -3.050 | -0.102 | **0.04** |  |  |  |  |
| Omicron BQ.1/CH.1.1/XBB.1.5:infection 3 | -1.811 | -3.199 | -0.422 | **0.01** |  |  |  |  |
| Time from most recent vaccination: Infection |  |  |  | **<0.001** |  |  |  |  |
| No vaccination:infection 2 | -0.563 | -1.119 | -0.006 | **0.04** |  |  |  |  |
| 14-90 days:infection 2 | 0.483 | 0.236 | 0.731 | **<0.001** |  |  |  |  |
| 90-180 days:infection 2 | -0.012 | -0.247 | 0.224 | 0.9 |  |  |  |  |
| No vaccination:infection 3 | -1.847 | -3.514 | -0.181 | **0.03** |  |  |  |  |
| 14-90 days:infection 3 | -0.397 | -1.166 | 0.372 | 0.3 |  |  |  |  |
| 90-180 days:infection 3 | -0.155 | -0.899 | 0.590 | 0.7 |  |  |  |  |

**Table S2. Effect estimates and 95% confidence intervals (95%CIs) in a normal linear regression model examining the association between Ct values and different characteristics.** The 95% CIs are calculated as estimates ± 1.96 × standard error of the estimates. Combined effects from interactions between variant, time from the most recent vaccination, and infection number are also shown in **Figure 1.** A separate model was built only including reinfections (second and third infections) and specifically examined the effects of Ct values and reported symptoms in the most recent previous infection (effects from Ct values are shown in **Figure S3**).

|  | Reinfection in Omicron BA.1 wave (27 December 2021 to 6 February 2022) | | | | Reinfection in Omicron BA.2 wave (14 March 2022 to 22 May 2022) | | | | | Reinfection in Omicron BA.4/5 wave (27 June 2022 to 6 November 2022) | | | | | Reinfection in Omicron BQ.1/CH.1.1/XBB.1.5 subvariants wave (7 November 2022 to 31 January 2023) | | | |
| --- | --- | --- | --- | --- | --- | --- | --- | --- | --- | --- | --- | --- | --- | --- | --- | --- | --- | --- |
|  | HR | 95%CI | | p-value | HR | 95%CI | | p-value | HR | | 95%CI | | p-value | HR | | 95%CI | | p-value |
| Sex (Male vs Female) | 0.91 | 0.83 | 0.99 | **0.03** | 0.85 | 0.79 | 0.91 | **<0.001** | 0.87 | | 0.83 | 0.90 | **<0.001** | 0.91 | | 0.88 | 0.95 | **<0.001** |
| Ethnicity (Non-white vs White) | 0.92 | 0.80 | 1.06 | 0.2 | 0.70 | 0.61 | 0.81 | **<0.001** | 0.88 | | 0.82 | 0.95 | **0.002** | 0.83 | | 0.76 | 0.90 | **<0.001** |
| Healthcare worker (Yes vs No) | 1.26 | 1.07 | 1.47 | **0.005** | 1.10 | 0.95 | 1.28 | 0.2 | 1.05 | | 0.96 | 1.13 | 0.3 | 1.12 | | 1.04 | 1.22 | **0.005** |
| Long-term health condition (Yes vs No) | 0.95 | 0.85 | 1.07 | 0.3 | 1.02 | 0.94 | 1.11 | 0.6 | 1.06 | | 1.01 | 1.11 | **0.02** | 1.03 | | 0.98 | 1.08 | 0.2 |
| Deprivation decile | 0.98 | 0.97 | 1.00 | **0.04** | 1.01 | 1.00 | 1.03 | 0.06 | 1.00 | | 0.99 | 1.00 | 0.4 | 1.00 | | 0.99 | 1.00 | 0.2 |
| Symptom in most recent previous infection (Classic vs No) | 0.92 | 0.82 | 1.04 | 0.2 | 0.87 | 0.79 | 0.96 | **0.007** | 1.04 | | 0.99 | 1.10 | 0.1 | 1.09 | | 1.03 | 1.14 | **0.003** |
| Symptom in most recent previous infection (Other vs No) | 1.04 | 0.91 | 1.19 | 0.5 | 1.02 | 0.91 | 1.13 | 0.6 | 1.03 | | 0.97 | 1.10 | 0.3 | 1.09 | | 1.03 | 1.16 | **0.005** |
| Any Ct<30 or LFD positive in previous infections | 1.00 | 0.79 | 1.27 | 0.9 | 0.81 | 0.67 | 0.97 | **0.03** | 1.01 | | 0.92 | 1.12 | 0.7 | 1.08 | | 0.98 | 1.18 | 0.1 |
| Region (Northern Ireland vs England) | 0.61 | 0.41 | 0.89 | **0.01** | 0.51 | 0.36 | 0.72 | **<0.001** | 0.93 | | 0.81 | 1.07 | 0.3 | 1.34 | | 1.19 | 1.50 | **<0.001** |
| Region (Scotland vs England) | 0.60 | 0.45 | 0.78 | **<0.001** | 0.80 | 0.67 | 0.96 | **0.02** | 0.97 | | 0.88 | 1.06 | 0.5 | 0.95 | | 0.87 | 1.04 | 0.2 |
| Region (Wales vs England) | 0.35 | 0.22 | 0.56 | **<0.001** | 0.88 | 0.71 | 1.08 | 0.2 | 1.01 | | 0.90 | 1.13 | 0.8 | 1.18 | | 1.07 | 1.29 | **<0.001** |
| Number of previous infections (2 vs 1) | 0.94 | 0.64 | 1.38 | 0.7 | 1.05 | 0.82 | 1.34 | 0.7 | 1.02 | | 0.94 | 1.11 | 0.6 | 1.01 | | 0.94 | 1.09 | 0.7 |
| Number of previous infections (3 vs 1) |  |  |  |  |  |  |  |  | 1.16 | | 0.70 | 1.93 | 0.6 | 1.31 | | 0.95 | 1.81 | **0.1** |
| Per 60 days after a most recent Pre-Alpha infection | 1.00 | 0.89 | 1.13 | 0.9 | 0.98 | 0.87 | 1.11 | 0.7 | 0.93 | | 0.79 | 1.10 | 0.4 | 0.88 | | 0.63 | 1.22 | 0.4 |
| Per 60 days after a most recent Alpha infection | 0.96 | 0.86 | 1.07 | 0.4 | 1.00 | 0.90 | 1.11 | 0.9 | 0.91 | | 0.84 | 0.98 | **0.009** | 0.86 | | 0.76 | 0.97 | **0.01** |
| Per 60 days after a most recent Delta infection | 0.75 | 0.61 | 0.91 | **0.004** | 1.14 | 1.07 | 1.21 | **<0.001** | 1.00 | | 0.97 | 1.04 | 0.9 | 0.89 | | 0.85 | 0.93 | **<0.001** |
| Per 60 days after a most recent Omicron BA.1 infection | |  |  |  |  |  |  |  | 1.18 | | 1.12 | 1.23 |  | 0.96 | | 0.91 | 1.01 | 0.1 |
| Per 60 days after a most recent Omicron BA.2 infection | |  |  |  |  |  |  |  | 1.31 | | 1.19 | 1.45 |  | 1.18 | | 1.12 | 1.24 | **<0.001** |
| Per 60 days after a most recent Omicron BA.4/5 infection | |  |  |  |  |  |  |  |  | |  |  |  | 1.44 | | 1.28 | 1.63 | **<0.001** |

**Table S3. Adjusted hazard ratios (HRs) with 95% CIs from parametric survival models examining the risk of reinfection in multiple Omicron infection waves (Omicron BA.1, BA.2, BA.4/5, and BQ.1/CH.1.1/XBB.1.5).** The 95% CIs are calculated by the exponent of the estimates ± 1.96 × standard error of the estimates. Effects of time from previous infection are shown in **Figure** **3,** from time from most recent vaccination in **Figure 4,** and of age and Ct values in **Figure S5, S6**. Effects from other covariates are shown in **Figure 2**. Results remain broadly similar in sensitivity analyses adjusted for background infection prevalence (differences in HR ranging from 0~0.02), except for small differences in the effect of age (shown in **Figure S5**) and region (shown in **Figure 2**).

| (A) Sensitivity analyses adjusted for infection prevalence | Omicron BA.1 wave (27 December 2021 to 6 February 2022) | | | | Omicron BA.2 wave (14 March 2022 to 22 May 2022) | | | | Omicron BA.4/5 wave (27 June 2022 to 6 November 2022) | | | | Omicron BQ.1/CH.1.1/XBB.1.5 subvariants wave (7 November 2022 to 31 January 2023) | | | |
| --- | --- | --- | --- | --- | --- | --- | --- | --- | --- | --- | --- | --- | --- | --- | --- | --- |
|  | HR | 95%CI | | p-value | HR | 95%CI | | p-value | HR | 95%CI | | p-value | HR | 95%CI | | p-value |
| Sex (Male vs Female) | 0.91 | 0.83 | 0.99 | **0.003** | 0.85 | 0.79 | 0.91 | **<0.001** | 0.87 | 0.83 | 0.90 | **<0.001** | 0.91 | 0.87 | 0.95 | **<0.001** |
| Ethnicity (Non-white vs White) | 0.92 | 0.79 | 1.06 | 0.2 | 0.71 | 0.61 | 0.81 | **<0.001** | 0.88 | 0.82 | 0.95 | **0.002** | 0.82 | 0.76 | 0.89 | **<0.001** |
| Healthcare worker (Yes vs No) | 1.26 | 1.08 | 1.48 | **0.003** | 1.10 | 0.95 | 1.28 | 0.2 | 1.05 | 0.96 | 1.13 | 0.3 | 1.17 | 1.08 | 1.26 | **<0.001** |
| Long-term health condition (Yes vs No) | 0.95 | 0.85 | 1.06 | 0.4 | 1.02 | 0.94 | 1.11 | 0.6 | 1.06 | 1.01 | 1.12 | **0.01** | 1.07 | 1.02 | 1.12 | **0.003** |
| Deprivation percentile | 0.98 | 0.97 | 1.00 | **0.04** | 1.01 | 1.00 | 1.03 | **0.03** | 1.00 | 0.99 | 1.00 | 0.4 | 0.99 | 0.99 | 1.00 | 0.1 |
| Symptom in most recent previous infection (Classic vs No) | 0.92 | 0.82 | 1.05 | 0.2 | 0.87 | 0.79 | 0.96 | **0.006** | 1.04 | 0.99 | 1.10 | 0.1 | 1.10 | 1.04 | 1.16 | **<0.001** |
| Symptom in most recent previous infection (Other vs No) | 1.04 | 0.91 | 1.19 | 0.5 | 1.02 | 0.91 | 1.13 | 0.7 | 1.03 | 0.97 | 1.10 | 0.4 | 1.10 | 1.03 | 1.17 | **0.003** |
| Any Ct<30 or LFD positive in all previous infections | 1.00 | 0.79 | 1.26 | 0.9 | 0.81 | 0.67 | 0.97 | **0.03** | 1.01 | 0.92 | 1.12 | 0.8 | 1.08 | 0.99 | 1.19 | 0.09 |
| Region (Northern Ireland vs England) | 0.58 | 0.39 | 0.85 | **0.005** | 0.64 | 0.45 | 0.91 | **0.01** | 0.94 | 0.82 | 1.08 | 0.4 | 1.10 | 0.98 | 1.24 | 0.1 |
| Region (Scotland vs England) | 0.75 | 0.56 | 0.98 | **0.04** | 0.77 | 0.64 | 0.93 | **0.006** | 0.98 | 0.89 | 1.07 | 0.6 | 0.99 | 0.91 | 1.08 | 0.7 |
| Region (Wales vs England) | 0.43 | 0.27 | 0.69 | **<0.001** | 0.86 | 0.70 | 1.06 | 0.2 | 0.96 | 0.86 | 1.07 | 0.4 | 1.04 | 0.95 | 1.15 | 0.4 |
| Number of previous infections (2 vs 1) | 0.93 | 0.64 | 1.37 | 0.7 | 1.06 | 0.83 | 1.35 | 0.6 | 1.03 | 0.94 | 1.11 | 0.6 | 1.01 | 0.94 | 1.08 | 0.7 |
| Number of previous infections (3 vs 1) |  |  |  |  |  |  |  |  | 1.16 | 0.70 | 1.93 | 0.6 | 1.39 | 1.02 | 1.89 | **0.04** |
| Per 60 days after a most recent Pre-Alpha infection | 1.00 | 0.89 | 1.13 | 0.9 | 0.98 | 0.87 | 1.11 | 0.8 | 0.94 | 0.80 | 1.10 | 0.4 | 0.88 | 0.63 | 1.22 | 0.4 |
| Per 60 days after a most recent Alpha infection | 0.95 | 0.85 | 1.06 | 0.4 | 1.00 | 0.90 | 1.11 | 0.9 | 0.91 | 0.84 | 0.98 | **0.009** | 0.85 | 0.76 | 0.95 | **0.006** |
| Per 60 days after a most recent Delta infection | 0.77 | 0.63 | 0.93 | **0.008** | 1.14 | 1.07 | 1.21 | **<0.001** | 1.00 | 0.97 | 1.04 | 0.9 | 0.89 | 0.85 | 0.93 | **<0.001** |
| Per 60 days after a most recent Omicron BA.1 infection | |  |  |  |  |  |  |  | 1.18 | 1.12 | 1.24 |  | 0.96 | 0.91 | 1.01 | 0.09 |
| Per 60 days after a most recent Omicron BA.2 infection | |  |  |  |  |  |  |  | 1.32 | 1.19 | 1.45 |  | 1.19 | 1.14 | 1.25 | **<0.001** |
| Per 60 days after a most recent Omicron BA.4/5 infection | | |  |  |  |  |  |  |  |  |  |  | 1.46 | 1.30 | 1.64 | **<0.001** |
| Prevalence (per 1% higher) | 1.20 | 1.14 | 1.26 | **<0.001** | 1.23 | 1.16 | 1.31 | **<0.001** | 1.33 | 1.26 | 1.41 | **<0.001** | 1.31 | 1.25 | 1.37 | **<0.001** |

| (B) Sensitivity analyses counting participants as being ‘at risk’ from the date of their first negative test | Omicron BA.1 wave (27 December 2021 to 6 February 2022) | | | | Omicron BA.2 wave (14 March 2022 to 22 May 2022) | | | | Omicron BA.4/5 wave (27 June 2022 to 6 November 2022) | | | | Omicron BQ.1/CH.1.1/XBB.1.5 subvariants wave (7 November 2022 to 31 January 2023) | | | |
| --- | --- | --- | --- | --- | --- | --- | --- | --- | --- | --- | --- | --- | --- | --- | --- | --- |
|  | HR | 95%CI | | p-value | HR | 95%CI | | p-value | HR | 95%CI | | p-value | HR | 95%CI | | p-value |
| Sex (Male vs Female) | 0.88 | 0.81 | 0.96 | **0.004** | 0.83 | 0.78 | 0.89 | **<0.001** | 0.86 | 0.83 | 0.90 | **<0.001** | 0.91 | 0.88 | 0.95 | **<0.001** |
| Ethnicity (Non-white vs White) | 0.93 | 0.81 | 1.07 | 0.3 | 0.71 | 0.63 | 0.81 | **<0.001** | 0.88 | 0.82 | 0.95 | **0.001** | 0.83 | 0.77 | 0.90 | **<0.001** |
| Healthcare worker (Yes vs No) | 1.25 | 1.07 | 1.46 | **0.004** | 1.16 | 1.02 | 1.32 | **0.02** | 1.04 | 0.96 | 1.13 | 0.4 | 1.13 | 1.04 | 1.22 | **0.003** |
| Long-term health condition (Yes vs No) | 0.95 | 0.86 | 1.06 | 0.4 | 1.04 | 0.96 | 1.12 | 0.3 | 1.08 | 1.03 | 1.13 | **0.002** | 1.04 | 0.99 | 1.09 | 0.2 |
| Deprivation percentile | 0.98 | 0.96 | 0.99 | **0.007** | 1.01 | 0.99 | 1.02 | 0.3 | 1.00 | 0.99 | 1.00 | 0.3 | 0.99 | 0.99 | 1.00 | 0.2 |
| Symptom in most recent previous infection (Classic vs No) | 0.89 | 0.80 | 1.00 | 0.06 | 0.85 | 0.78 | 0.93 | **<0.001** | 1.03 | 0.97 | 1.08 | 0.4 | 1.08 | 1.02 | 1.14 | **0.004** |
| Symptom in most recent previous infection (Other vs No) | 1.00 | 0.88 | 1.14 | 0.9 | 1.01 | 0.91 | 1.11 | 0.8 | 1.02 | 0.96 | 1.08 | 0.6 | 1.09 | 1.02 | 1.16 | **0.007** |
| Any Ct<30 or LFD positive in all previous infections | 1.00 | 0.80 | 1.25 | 0.9 | 0.89 | 0.75 | 1.04 | 0.1 | 1.01 | 0.91 | 1.11 | 0.8 | 1.09 | 0.99 | 1.19 | 0.07 |
| Region (Northern Ireland vs England) | 0.58 | 0.40 | 0.85 | **0.005** | 0.45 | 0.33 | 0.63 | **<0.001** | 0.94 | 0.82 | 1.08 | 0.4 | 1.31 | 1.17 | 1.47 | **<0.001** |
| Region (Scotland vs England) | 0.57 | 0.44 | 0.74 | **<0.001** | 0.80 | 0.67 | 0.94 | **0.007** | 0.94 | 0.86 | 1.03 | 0.2 | 0.96 | 0.88 | 1.04 | 0.3 |
| Region (Wales vs England) | 0.37 | 0.24 | 0.56 | **<0.001** | 0.80 | 0.66 | 0.98 | **0.03** | 0.97 | 0.87 | 1.08 | 0.6 | 1.16 | 1.06 | 1.28 | **0.002** |
| Number of previous infections (2 vs 1) | 0.87 | 0.62 | 1.24 | 0.4 | 0.92 | 0.76 | 1.11 | 0.4 | 1.02 | 0.94 | 1.11 | 0.6 | 1.00 | 0.93 | 1.07 | 0.9 |
| Number of previous infections (3 vs 1) |  |  |  |  |  |  |  |  | 1.23 | 0.77 | 1.95 | 0.4 | 1.19 | 0.87 | 1.63 | 0.3 |
| Per 60 days after a most recent Pre-Alpha infection | 0.99 | 0.89 | 1.12 | 0.9 | 0.97 | 0.86 | 1.09 | 0.6 | 0.92 | 0.78 | 1.08 | 0.3 | 0.88 | 0.63 | 1.23 | 0.5 |
| Per 60 days after a most recent Alpha infection | 0.95 | 0.85 | 1.06 | 0.4 | 0.99 | 0.89 | 1.10 | 0.8 | 0.90 | 0.83 | 0.96 | **0.004** | 0.86 | 0.76 | 0.97 | **0.01** |
| Per 60 days after a most recent Delta infection | 1.46 | 1.35 | 1.58 | **<0.001** | 1.09 | 1.04 | 1.15 | **<0.001** | 1.00 | 0.96 | 1.03 | 0.8 | 0.89 | 0.85 | 0.93 | **<0.001** |
| Per 60 days after a most recent Omicron BA.1 infection | |  |  |  | 2.29 | 1.93 | 2.72 | **<0.001** | 1.24 | 1.19 | 1.30 | **<0.001** | 0.96 | 0.91 | 1.01 | 0.1 |
| Per 60 days after a most recent Omicron BA.2 infection | |  |  |  |  |  |  |  | 1.75 | 1.64 | 1.88 | **<0.001** | 1.23 | 1.18 | 1.29 | **<0.001** |
| Per 60 days after a most recent Omicron BA.4/5 infection | | |  |  |  |  |  |  | 3.61 | 2.47 | 5.29 | **<0.001** | 1.71 | 1.58 | 1.85 | **<0.001** |

| (C) Sensitivity analyses only including infections defined by positive test results from the study | Reinfection in Omicron BA.1 wave (27 December 2021 to 6 February 2022) | | | | Reinfection in Omicron BA.2 wave (14 March 2022 to 22 May 2022) | | | | | Reinfection in Omicron BA.4/5 wave (27 June 2022 to 6 November 2022) | | | | | Reinfection in Omicron BQ.1/CH.1.1/XBB.1.5 subvariants wave (7 November 2022 to 31 January 2023) | | | |
| --- | --- | --- | --- | --- | --- | --- | --- | --- | --- | --- | --- | --- | --- | --- | --- | --- | --- | --- |
|  | HR | 95%CI | | p-value | HR | 95%CI | | p-value | HR | | 95%CI | | p-value | HR | | 95%CI | | p-value |
| Sex (Male vs Female) | 0.95 | 0.76 | 1.19 | 0.6 | 0.87 | 0.75 | 1.00 | 0.05 | 0.87 | | 0.81 | 0.93 | **<0.001** | 0.93 | | 0.87 | 0.99 | **0.02** |
| Ethnicity (Non-white vs White) | 1.17 | 0.80 | 1.70 | 0.4 | 0.92 | 0.69 | 1.22 | 0.6 | 0.95 | | 0.82 | 1.10 | 0.5 | 0.89 | | 0.77 | 1.01 | 0.08 |
| Healthcare worker (Yes vs No) | 0.97 | 0.59 | 1.59 | 0.9 | 0.91 | 0.64 | 1.29 | 0.6 | 0.91 | | 0.77 | 1.09 | 0.3 | 1.07 | | 0.93 | 1.24 | 0.3 |
| Long-term health condition (Yes vs No) | 0.66 | 0.49 | 0.89 | **0.007** | 0.92 | 0.77 | 1.09 | 0.3 | 1.00 | | 0.91 | 1.09 | 0.9 | 1.01 | | 0.93 | 1.08 | 0.9 |
| Deprivation decile | 0.98 | 0.94 | 1.01 | 0.3 | 1.02 | 0.99 | 1.05 | 0.2 | 1.00 | | 0.99 | 1.01 | 0.9 | 1.00 | | 0.98 | 1.01 | 0.5 |
| Symptom in most recent previous infection (Classic vs No) | 0.95 | 0.72 | 1.25 | 0.7 | 0.95 | 0.80 | 1.13 | 0.5 | 1.01 | | 0.92 | 1.09 | 0.9 | 1.07 | | 1.00 | 1.16 | 0.06 |
| Symptom in most recent previous infection (Other vs No) | 0.82 | 0.57 | 1.17 | 0.3 | 0.82 | 0.65 | 1.03 | 0.1 | 1.04 | | 0.93 | 1.16 | 0.5 | 1.06 | | 0.95 | 1.17 | 0.3 |
| Any Ct<30 or LFD positive in previous infections | 1.09 | 0.63 | 1.90 | 0.8 | 0.74 | 0.51 | 1.07 | 0.1 | 0.97 | | 0.82 | 1.15 | 0.7 | 1.04 | | 0.89 | 1.22 | 0.6 |
| Region (Northern Ireland vs England) | 1.96 | 1.30 | 2.97 | **0.001** | 0.95 | 0.67 | 1.36 | 0.8 | 1.28 | | 1.11 | 1.48 | **<0.001** | 1.58 | | 1.40 | 1.78 | **<0.001** |
| Region (Scotland vs England) | 1.86 | 1.36 | 2.56 | **<0.001** | 1.45 | 1.18 | 1.78 | **<0.001** | 1.33 | | 1.20 | 1.47 | **<0.001** | 1.12 | | 1.02 | 1.22 | **0.02** |
| Region (Wales vs England) | 1.11 | 0.68 | 1.82 | 0.7 | 1.61 | 1.28 | 2.03 | **<0.001** | 1.39 | | 1.23 | 156 | **<0.001** | 1.38 | | 1.25 | 1.53 | **<0.001** |
| Number of previous infections (2 vs 1) | 0.67 | 0.16 | 2.76 | 0.6 | 0.94 | 0.48 | 1.82 | 0.7 | 1.14 | | 0.94 | 1.39 | 0.07 | 1.15 | | 1.00 | 1.32 | 0.06 |
| Number of previous infections (3 vs 1) |  |  |  |  |  |  |  |  | 2.27 | | 0.94 | 5.46 | 0.6 | 2.16 | | 1.30 | 3.60 | **0.003** |
| Per 60 days after a most recent Pre-Alpha infection | 0.95 | 0.71 | 1.26 | 0.7 | 0.93 | 0.76 | 1.16 | 0.5 | 1.07 | | 0.81 | 1.41 | 0.6 | 0.58 | | 0.36 | 0.93 | 0.02 |
| Per 60 days after a most recent Alpha infection | 0.91 | 0.70 | 1.18 | 0.5 | 0.92 | 0.77 | 1.11 | 0.4 | 0.89 | | 0.79 | 1.01 | 0.06 | 0.86 | | 0.72 | 1.04 | **0.1** |
| Per 60 days after a most recent Delta infection | 0.88 | 0.53 | 1.47 | 0.6 | 1.19 | 1.04 | 1.36 | **0.01** | 1.01 | | 0.93 | 1.09 | 0.9 | 0.89 | | 0.82 | 0.98 | **0.01** |
| Per 60 days after a most recent Omicron BA.1 infection | |  |  |  |  |  |  |  | 1.16 | | 1.06 | 1.27 | **0.001** | 0.90 | | 0.82 | 0.98 | **0.02** |
| Per 60 days after a most recent Omicron BA.2 infection | |  |  |  |  |  |  |  | 1.29 | | 1.11 | 1.49 | **<0.001** | 1.18 | | 1.10 | 1.27 | **<0.001** |
| Per 60 days after a most recent Omicron BA.4/5 infection | |  |  |  |  |  |  |  |  | |  |  |  | 1.42 | | 1.22 | 1.66 | **<0.001** |

**Table S4. Sensitivity analyses estimating adjusted hazard ratios (HRs) with 95% CIs in parametric survival models examining the risk of reinfection in multiple Omicron infection waves (Omicron BA.1, BA.2, BA.4/5, and BQ.1/CH.1.1/XBB.1.5).** The 95% CIs are calculated as the exponent of the estimates ± 1.96 × standard error of the estimates. Table A shows the results in models adjusted for background infection prevalence. Table B shows the results in models counting participants as being ‘at risk’ from the date of their first negative PCR test in the study following each infection rather than 120 days after their previous infection. Results remain broadly similar to the primary analysis. Table C shows the results in models only including infections defined by positive test results from the study (so not influenced by test seeking behaviour).

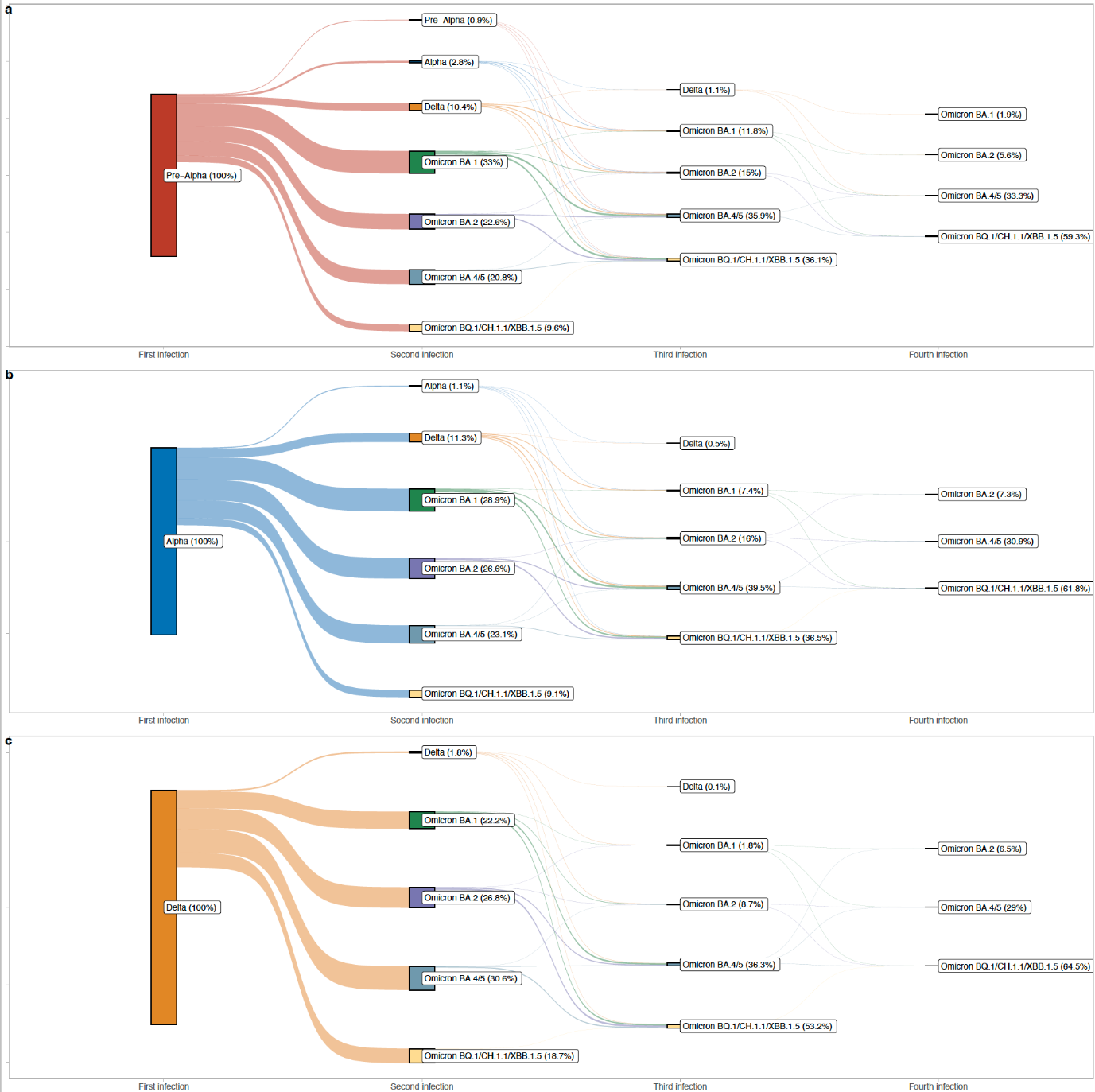

**Figure S1. Percentages of with first, second, third, and fourth infection by variants of the first infection (Pre-Alpha, Alpha, and Delta) and subsequent infections.**

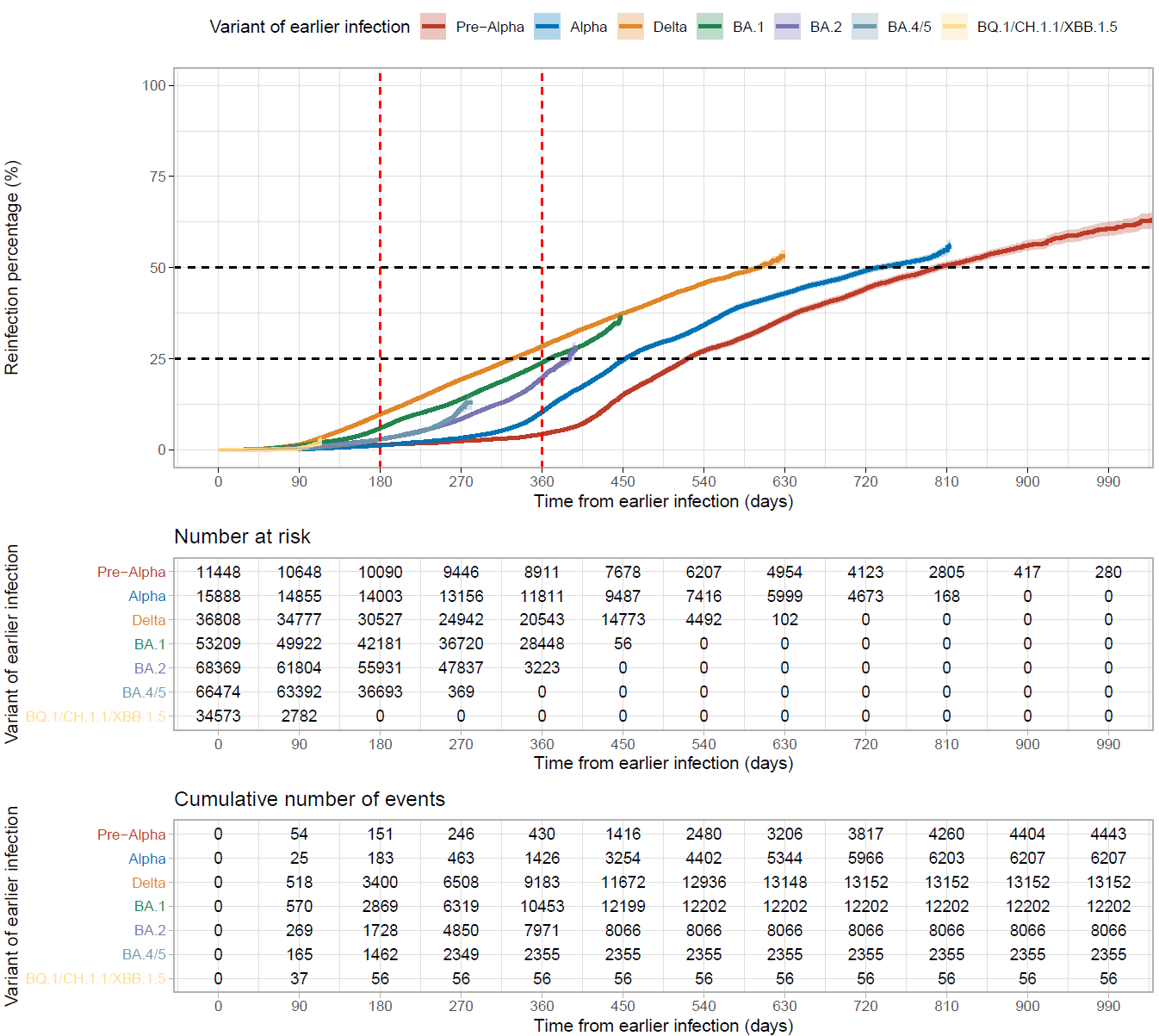

**Figure S2. Unadjusted percentage reinfected over time from the earlier infection by variant of earlier infection using Kaplan-Meier estimation.** Survival models were built from the start of the earlier infection and censored at each participant’s last visit date. Black horizontal dashed lines show the time to 25% and 50% of the subgroup becoming reinfected. Red vertical dashed lines show the percentage reinfected 6 and 12 months after the earlier infection. Number at risk and cumulative number of events are shown in tables by 90 days. 95% CIs are calculated as estimates ± 1.96 × standard error of the estimates.

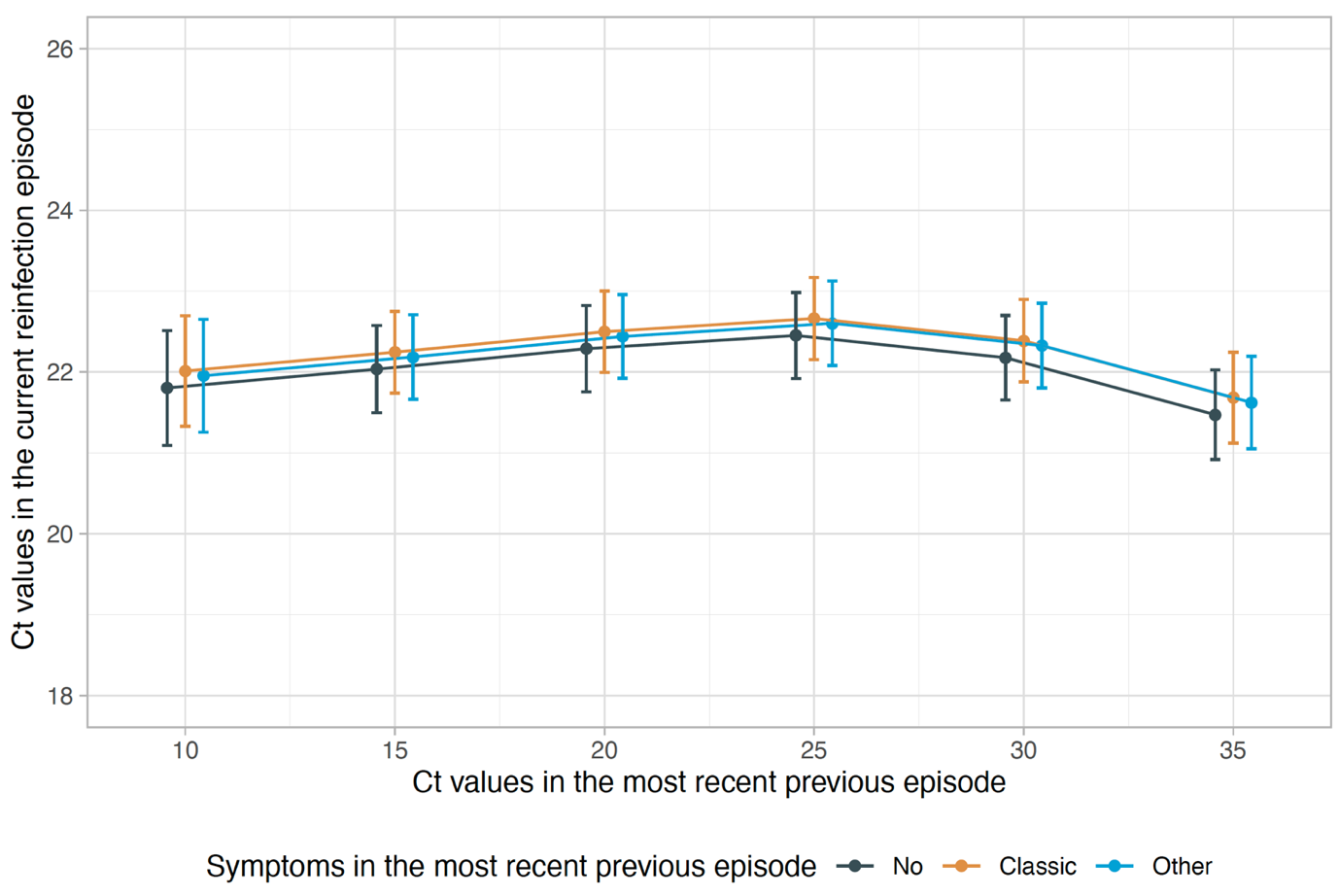

**Figure S3. Association between Ct values in the current reinfection with Ct values and symptoms in the most recent previous infection.** The 95% CIs are calculated as estimates ± 1.96 × standard error of the estimates. Adjusted (**Table S2**) for age, sex, ethnicity, reporting working in healthcare, reporting having a long-term health condition, deprivation percentile, reported symptoms in this infection, number of positive tests in the infection, infection variant, time from most recent vaccination.

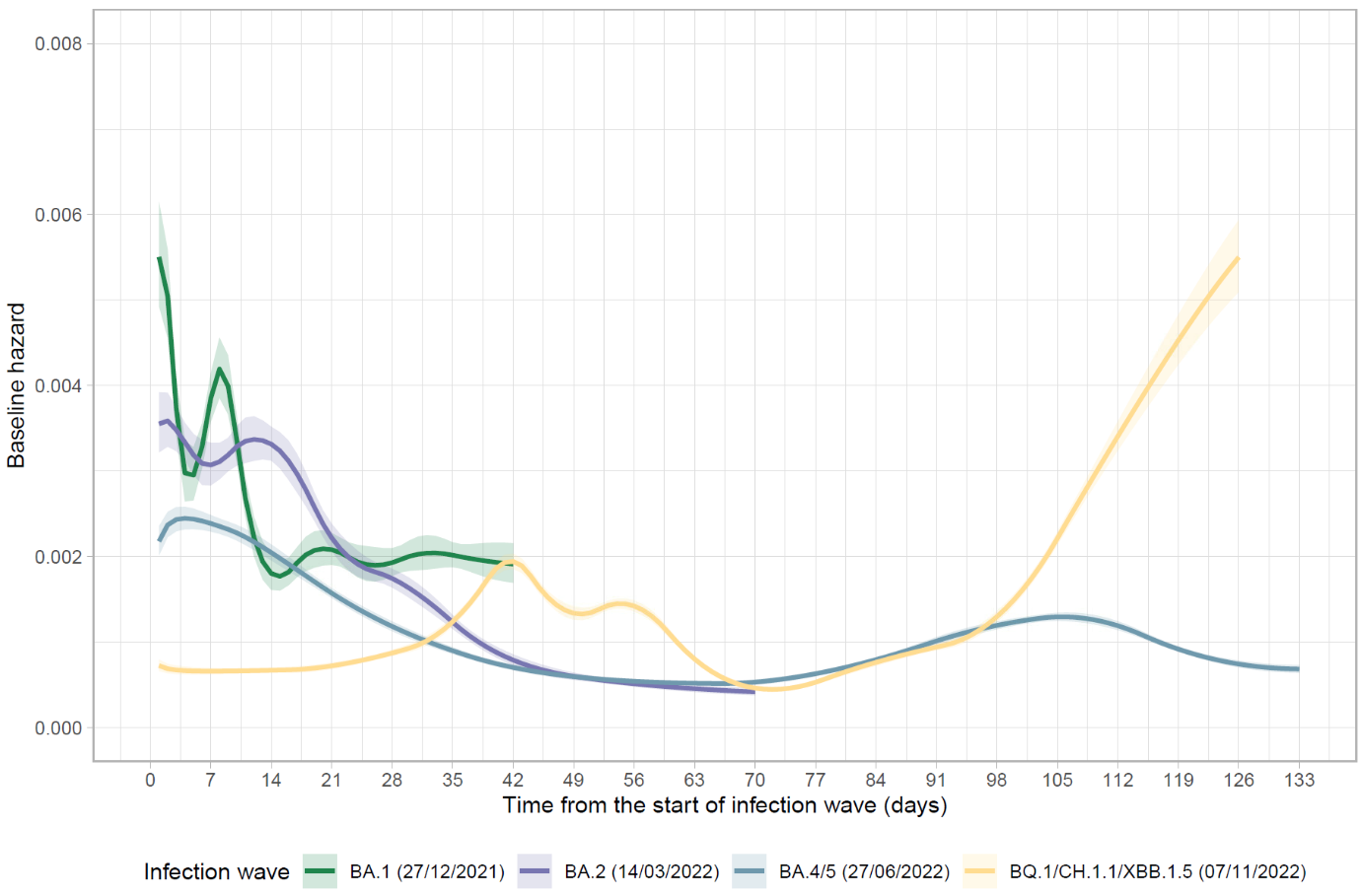

**Figure S4. Baseline reinfection risk over calendar time from the start of the BA.1, BA.2, BA.4/5, and BQ.1/CH.1.1/XBB.1.5 infection waves estimated using unadjusted flexible parametric survival models.** 95% CIs are calculated as estimates ± 1.96 × standard error of the estimates.

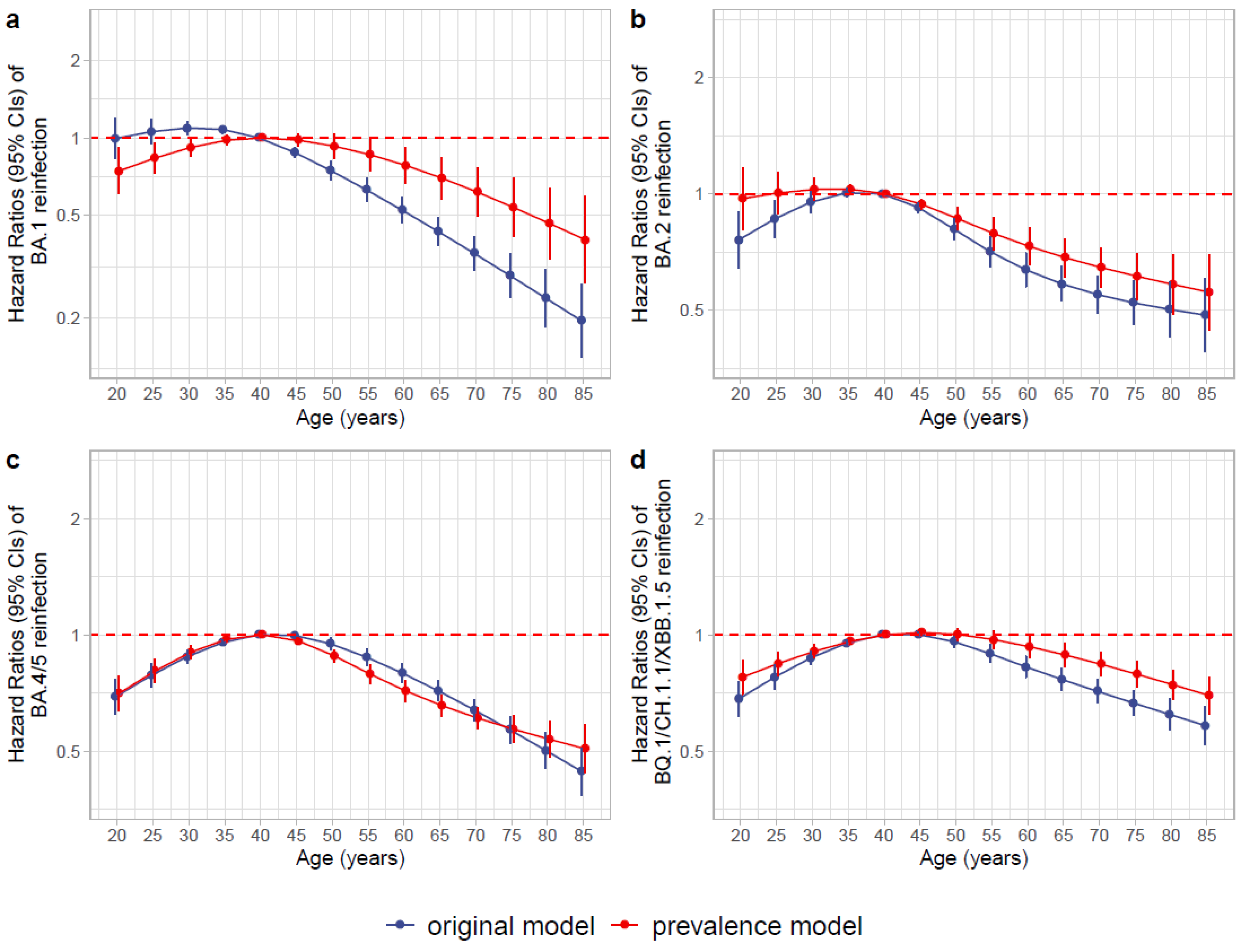

**Figure S5. Independent association between age (Hazard Ratios with 95% CIs) and risk of reinfection in the Omicron BA.1, BA.2, BA.4/5, and BQ.1/CH.1.1/XBB.1.5 waves.** The 95% CIs are calculated as the exponent of estimates ± 1.96 × standard error of the estimates. The differences in hazard ratios by age between the original model and the model adjusted for background infection prevalence generally show slight attenuation towards the null adjusting for background prevalence. See **Tables S3 and S4** for other factors.

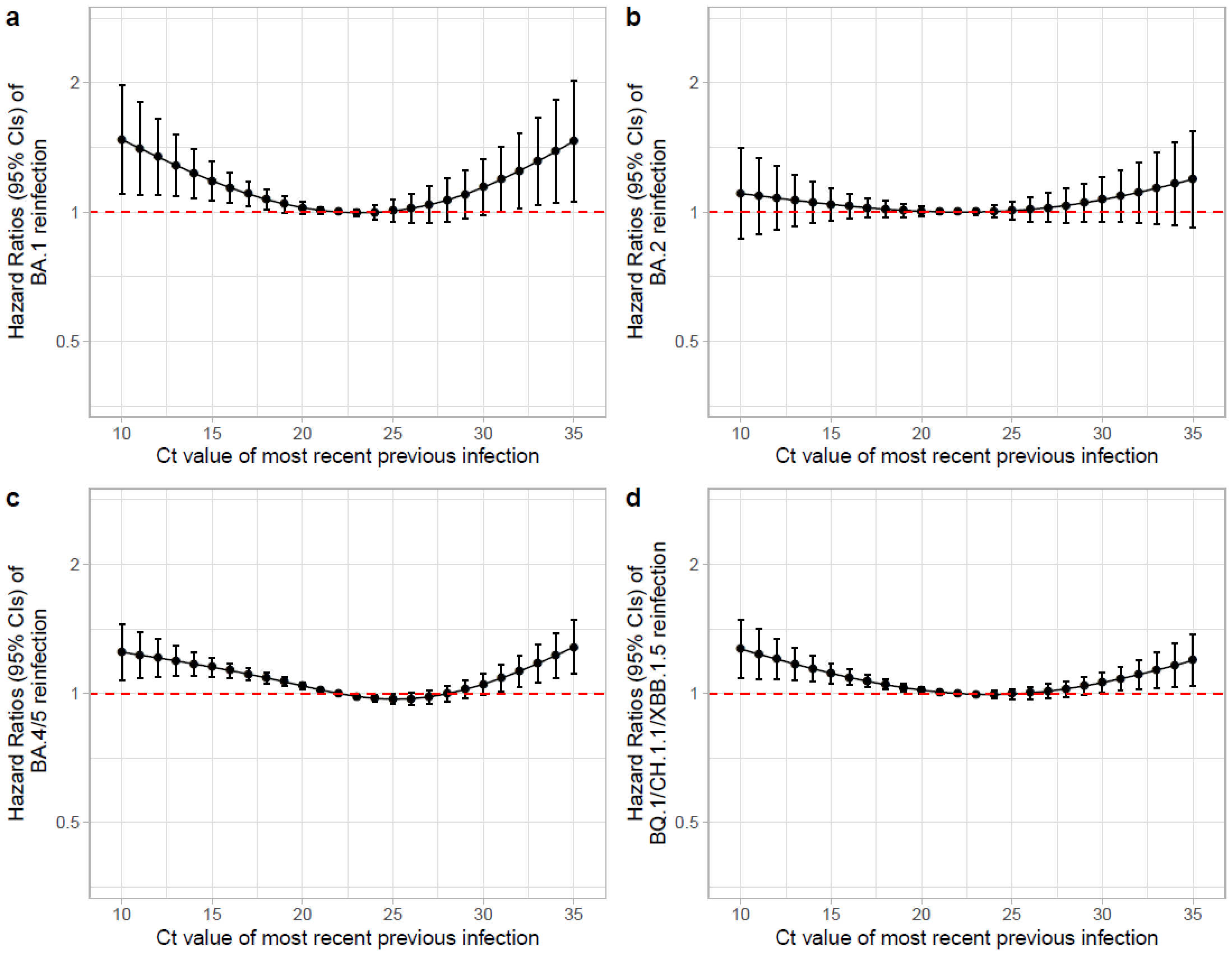

**Figure S6. Independent association between Ct value of the previous infection and risk (Hazard Ratios with 95% CIs) of reinfection in the Omicron BA.1, BA.2, BA.4/5, and BQ.1/CH.1.1/XBB.1.5 waves.** The 95% CIs are calculated as the exponent of estimates ± 1.96 × standard error of the estimates. Results remain unchanged in sensitivity analyses adjusted for background infection prevalence. See **Tables S3 and S4** for other factors.

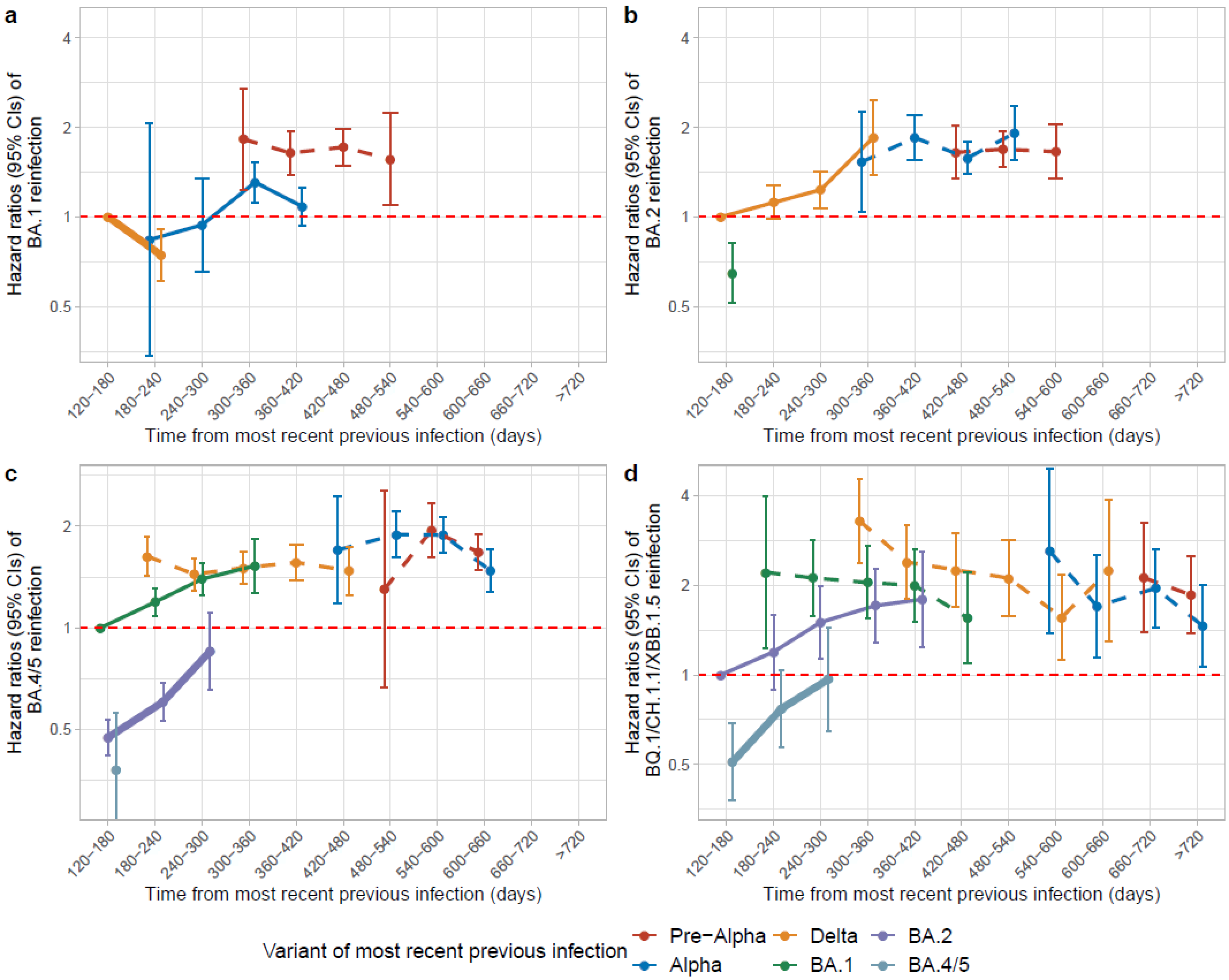

**Figure S7. Risk (Hazard Ratios with 95% CIs) of reinfections during Omicron BA.1 (a), Omicron BA.2 (b), Omicron BA.4/5 (c) and Omicron BQ.1/CH.1.1/XBB.1.5 (d) waves by time from most recent previous infection and variant of most recent previous infection.** Time from previous infection was categorised as 120-180, 180-240, 240-300, 300-360, 360-420, 420-480, 480-540, 540-600, 600-660, 660-720, >720 days and its effect modelled categorically (**Figure 3** shows results modelling the categories as a linear trend). Risk is presented versus a reference category of 120-180 days from an infection in the wave starting ~6 months before the current wave (Delta for BA.1 and BA.2 waves, BA.1 for BA.4/5 waves and BA.2 for BQ.1/CH.1.1/XBB.1.5 waves). Line type and width represent the sequence of variants for better comparisons across waves (thick solid line represents the previous variant, thin solid line represents the penultimate variant, and dashed lines represent earlier variants). The 95% CIs are calculated as the exponent of estimates ± 1.96 × standard error of the estimates. Adjusted for age, sex, ethnicity, reporting working in healthcare, reporting having a long-term health condition, deprivation percentile, infection variant, time from most recent vaccination, region, number of previous infections, symptoms in most recent infection and whether any previous infection had Ct<30 or was LFD positive. Results remain similar in sensitivity analyses adjusted for background infection prevalence.

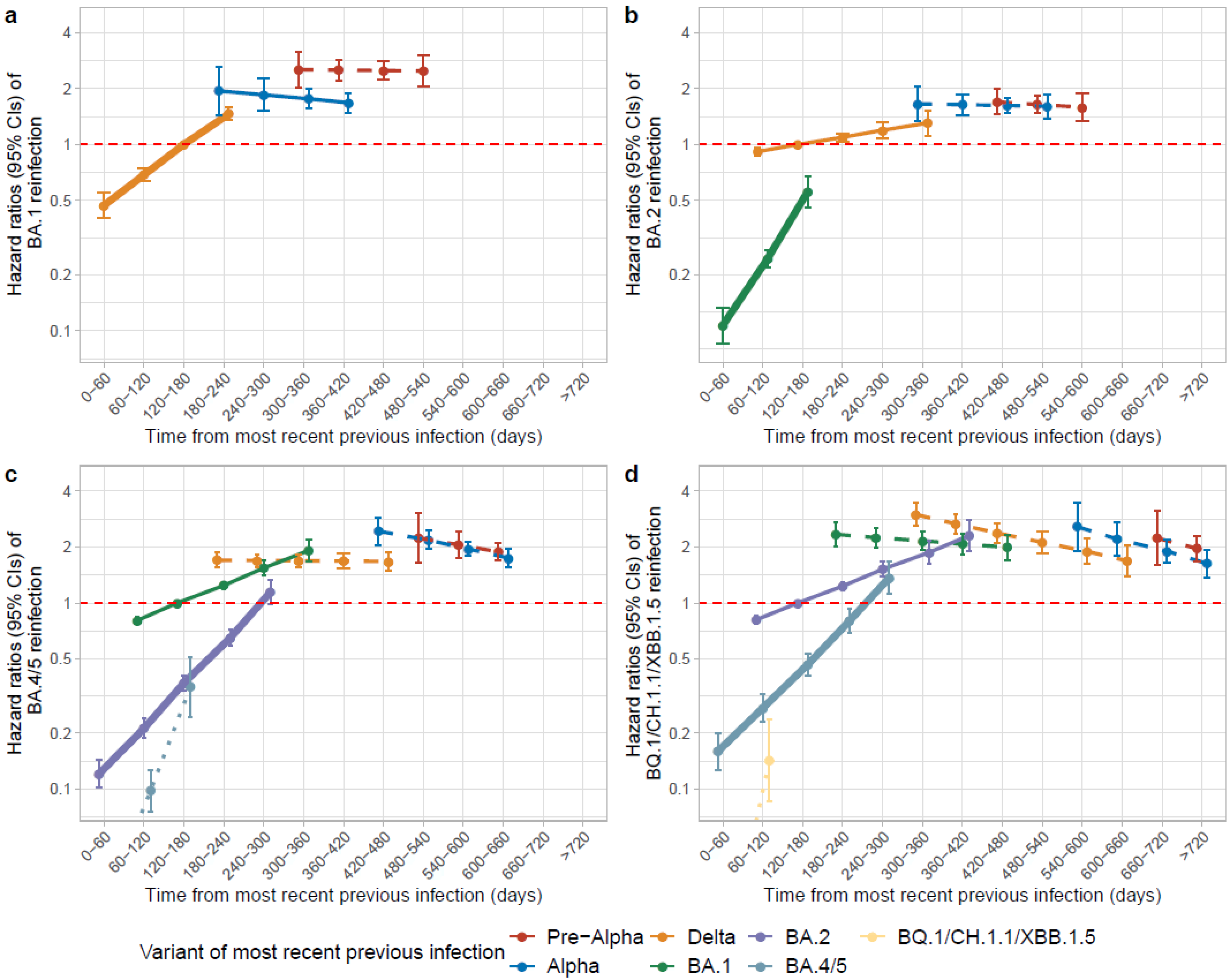

**Figure S8. Risk (Hazard Ratios with 95% CIs) of reinfections during the Omicron BA.1 (a), Omicron BA.2 (b), Omicron BA.4/5 (c) and Omicron BQ.1/CH.1.1/XBB.1.5 (d) waves by time from most recent previous infection and variant of most recent previous infection in sensitivity analysis counting participants as being ‘at risk’ from the date of their first negative study PCR test following the infection.** Time from previous infection was categorised as 0-60, 60-120, 120-180, 180-240, 240-300, 300-360, 360-420, 420-480, 480-540, 540-600, 600-660, 660-720, >720 days and its effect modelled as a trend over these categories (see **Figure S9** for categorical effects). Risk is presented versus a reference category of 120-180 days from an infection in the wave starting ~6 months before the current wave (Delta for BA.1 and BA.2 waves, BA.1 for BA.4/5 waves and BA.2 for BQ.1/CH.1.1/XBB.1.5 waves). Line type and width represent the sequence of variants for better comparisons across waves (thick solid line represents the previous variant, thin solid line represents the penultimate variant, and dashed lines represent earlier variants). The 95% CIs are calculated as exponent of estimates ± 1.96 × standard error of the estimates. Adjusted for age, sex, ethnicity, reporting working in healthcare, reporting having a long-term health condition, deprivation percentile, infection variant, time from most recent vaccination, region, number of previous infections, symptoms in most recent infection and whether any previous infection had Ct<30 or was LFD positive.

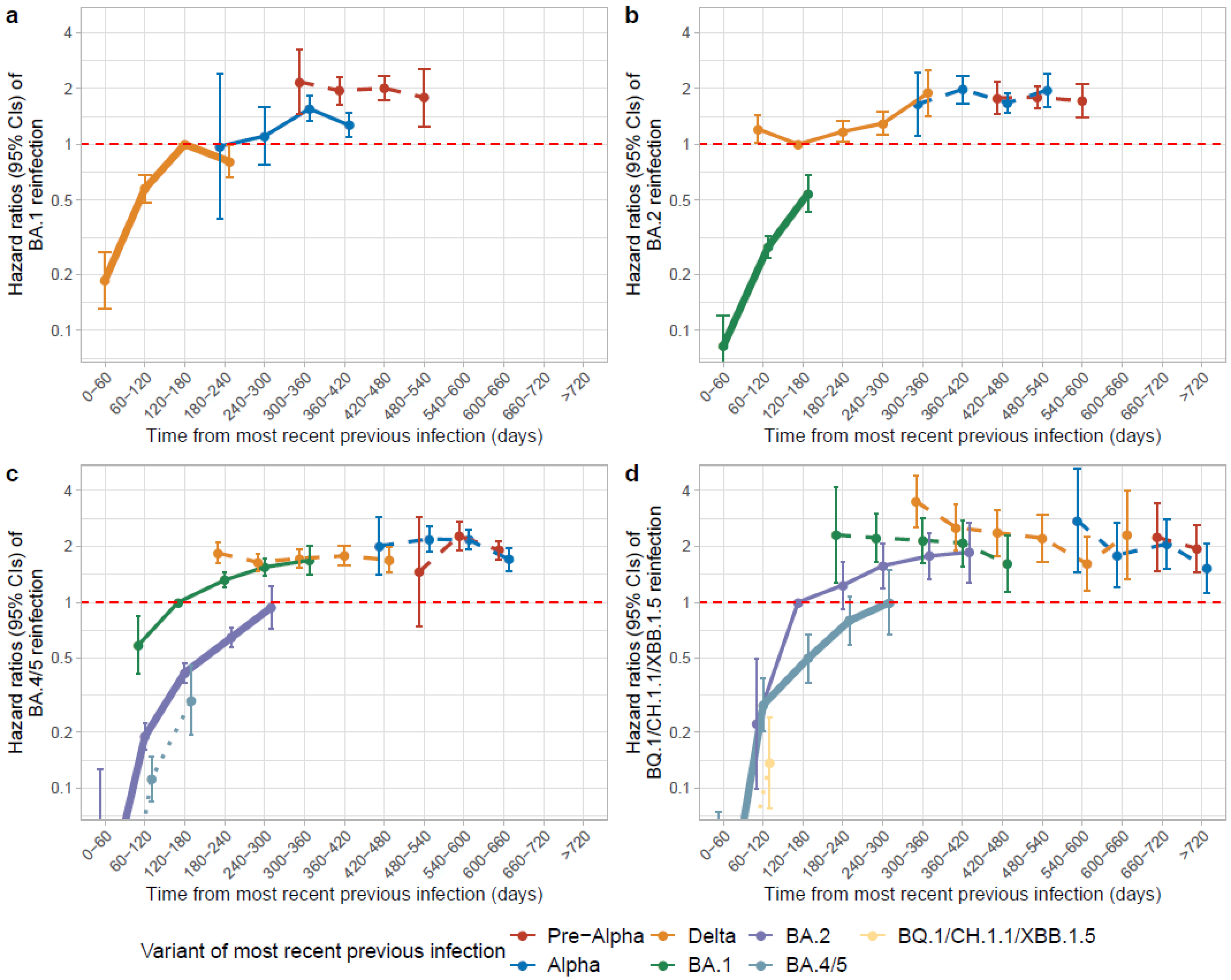

**Figure S9. Risk (Hazard Ratios with 95% CIs) of reinfections during Omicron BA.1 (a), Omicron BA.2 (b), Omicron BA.4/5 (c) and Omicron BQ.1/CH.1.1/XBB.1.5 (d) waves by time from most recent previous infection and variant of most recent previous infection in sensitivity analyses counting participants as being ‘at risk’ from the date of their first negative study PCR test following the infection.** Time from previous infection was categorised as 0-60, 60-120, 120-180, 180-240, 240-300, 300-360, 360-420, 420-480, 480-540, 540-600, 600-660, 660-720, >720 days and its effect modelled categorically (**Figure S8** shows results modelling the categories as a linear trend). Risk is presented versus a reference category of 120-180 days from an infection in the wave starting ~6 months before the current wave (Delta for BA.1 and BA.2 waves, BA.1 for BA.4/5 waves and BA.2 for BQ.1/CH.1.1/XBB.1.5 waves). Line type and width represent the sequence of variants for better comparisons across waves (thick solid line represents the previous variant, thin solid line represents the penultimate variant, and dashed lines represent earlier variants). The 95% CIs are calculated as the exponent of estimates ± 1.96 × standard error of the estimates. Adjusted for age, sex, ethnicity, reporting working in healthcare, reporting having a long-term health condition, deprivation percentile, infection variant, time from most recent vaccination, region, number of previous infections, symptoms in most recent infection and whether any previous infection had Ct<30 or was LFD positive.
